# Integrated antigenic and nucleic acid detection in single virions and virion-infected host-derived extracellular vesicles

**DOI:** 10.1101/2023.08.31.23292825

**Authors:** Kim Truc Nguyen, Xilal Y. Rima, Luong T. H. Nguyen, Xinyu Wang, Kwang Joo Kwak, Min Jin Yoon, Hong Li, Chi-Ling Chiang, Jacob Doon-Ralls, Kelsey Scherler, Shannon Fallen, Stephanie L. Godfrey, Julie A. Wallick, Setty M. Magaña, Andre F. Palmer, Inyoul Lee, Christopher C. Nunn, Kimberly M. Reeves, Henry G. Kaplan, Jason D. Goldman, James R. Heath, Kai Wang, Preeti Pancholi, L. James Lee, Eduardo Reátegui

**Affiliations:** William G. Lowrie Department of Chemical and Biomolecular Engineering, The Ohio State University; Columbus, OH 43210, USA; Diabetes and Metabolism Research Center, The Ohio State University Wexner Medical Center, Columbus; OH, 43210, United States; Spot Biosystems Ltd.; Palo Alto, CA 94301, USA; Institute for Systems Biology; Seattle, WA 98109, USA; Providence Swedish Medical Center; Seattle, WA 98104, USA; Translational Neuroimmunology, Center for Clinical and Translational Research, Nationwide Children’s Hospital; Columbus, OH 43205, USA; Providence Swedish Cancer Institute; Seattle, WA 98104, USA; Division of Allergy and Infectious Diseases, University of Washington; Seattle, WA 98195, USA; Department of Pathology, The Ohio State University Wexner Medical Center; Columbus, OH 43203, USA; Comprehensive Cancer Center, The Ohio State University; Columbus, OH 43210, USA

**Author notes:** Authors contributed equally (Kim Truc Nguyen, Xilal Y. Rima).

**Keywords:** Single virion, SARS-CoV-2, COVID-19, single extracellular vesicle, multiparametric

## Abstract

Virion-mediated outbreaks are imminent and despite rapid responses, they continue to cause adverse symptoms and death. Therefore, tunable, sensitive, high-throughput assays are needed to control future virion-mediated outbreaks. Herein, we developed a tunable *in situ* assay to selectively sort virions and infected host-derived extracellular vesicles (IHD-EVs) and simultaneously detect antigens and nucleic acids at a single-particle resolution. The Biochip Antigen and RNA Assay (BARA) enhanced sensitivities, enabling the detection of virions in asymptomatic patients, genetic mutations in single virions, and the continued long-term expression of virion-RNA in the IHD-EVs of post-acute sequelae of COVID-19 patients. The BARA revealed highly accurate diagnoses by simultaneously detecting the spike glycoprotein and nucleocapsid-encoding RNA on single SARS-CoV-2 virions in saliva and nasopharyngeal swab samples. Altogether, the single-particle detection of antigens and virion-RNA provides a tunable framework for the diagnosis, monitoring, and mutation screening of current and future outbreaks.

**Teaser:** The BARA enables antigenic and nucleic acid testing in single virions for unprecedented perspectives on viral diseases

## Introduction

The emergence of infectious diseases is rising and is dominated by zoonoses (1), which are the transmission of pathogens from animals to humans that originate via a myriad of interspecies interactions (2). Human history is concomitant with zoonoses, begetting pandemics, epidemics, and endemics that have plagued the human experience, the former two requiring interpersonal transmission and the latter typically contained in the individual (3). Although we have coexisted with zoonotic pathogens, their outbreaks continue to disrupt the social fabric at an individual level, such as inducing psychological distress, or at the societal level, such as burdening the economy (4–7). The coronavirus disease of 2019 (COVID-19) caused by the severe acute respiratory syndrome coronavirus 2 (SARS-CoV-2) is no exception, resulting in the reported infection of 767 million people and the deaths of 6.9 million worldwide to date (8). While the number of deaths is staggering and continues to increase, in the United States of America (USA), excess deaths were disproportionately higher for Black, Latino, and American Indian/Alaska Native persons, exacerbating racial inequities across the country (9). With global warming and human land use encouraging the interaction of species via habitat reduction, the number of zoonoses is expected to increase (10, 11). Therefore, methods to slow the transmission of zoonoses via rapidly tunable diagnostic assays that provide highly sensitive detection, molecular subtyping, and follow-up monitoring of pathogens are necessary to mitigate future epidemics via containment measures.

Two prevalent detection methods for the diagnosis of zoonoses are nucleic-acid-based and antigen-based technologies (12). While highly sensitive, nucleic-acid-based technologies are limited at detecting genetic mutations, novel zoonoses, or low-virion counts; whereas, antigen-based technologies require post-acute immune responses (12–16). Quantitative reverse-transcriptase polymerase chain reaction (qRT-PCR), a nucleic-acid-based assay, was the primary diagnostic utilized to combat the transmission of the SARS-CoV-2 virion despite its false-negative results and requirement for laboratory equipment and reagents (17, 18). On the other hand, rapid-antigen tests that target intrinsic virion proteins granted accessibility to facile COVID-19 testing for the general public, but are less sensitive than qRT-PCR (19). Bulk-analysis diagnostics are subject to the dilution of virions within the biofluid, which can be averted by detecting the biomolecular expression of single virions (20). Novel single-virion technologies have emerged as a response to the shortcomings of traditional diagnostics to enhance sensitivities and specificities (21–24). However, the simultaneous detection of antigens and nucleic acids in single virions to further improve detection limits is yet to be realized. Virions are biogenetically and morphologically similar to extracellular vesicles (EVs) (25), which are cell-derived lipid nanoparticles containing bioactive molecules (26). The advent of research in EVs has sought to characterize biomolecular signatures to unravel their vast heterogeneity (27), which has promoted the engineering of *in situ* single-EV technologies (28–36). Coupling *in situ* labeling with high-resolution microscopy, such as total internal reflection fluorescence microscopy (TIRFM), has further enabled the colocalization of proteins and nucleic acids in single EVs (37). TIRFM has been utilized to investigate single-virion dynamics upon entry or release from the plasma membrane (38–40). Therefore, the translation of single-EV technologies to single virions seems a natural transition to screen for both the presence of virions and infected tissue. Combining TIRFM and single-EV labeling techniques may provide a unique perspective into single-virion biomolecular signatures via colocalization of antigenic and nucleic acid detection.

Herein we present the Biochip Antigen and RNA Assay (BARA), which isolates single virions and virion-infected host-derived EVs (IHD-EVs) from complex biofluids via positive immunoselection and infection mechanisms. The BARA combines immunofluorescence (IF) and fluorescent *in situ* hybridization (FISH) with TIRFM providing high-resolution qualitative and quantitative antigenic and nucleic acid expression of single particles. The BARA was validated with the SARS-CoV-2 virion following the guidelines for Emergency Use Authorization (EUA) regulated by the United States Food and Drug Administration (FDA). By progressing toward single-virion detection, the BARA outperformed quantitative reverse transcription polymerase chain reaction (qRT-PCR) by one order of magnitude regarding the limit of detection (LoD), which upon combining antigenic and nucleic acid detection, yielded sensitivities of 100 % and 95 % and specificities of 100 % and 100 % for saliva and nasopharyngeal swab (NS) samples, respectively. Furthermore, the BARA revealed the continued long-term expression of virion-RNA in IHD-EVs from post-acute sequelae of COVID-19 (PASC) patient plasma. The success of the work provides a tunable framework to interrogate single virions and long-term infections via the simultaneous detection of biomolecules in single particles, which can be easily adapted by customizing the antibodies and proteins for immunoselection and the probes for their subsequent detection.

## Results

### Simultaneous detection of biomolecules on intact single virions and EVs

The BARA is a high-throughput assay that multiplexes signals from antigens and nucleic acids utilizing IF and FISH on single virions and IHD-EVs derived from complex biofluids, such as blood plasma, saliva, and NS. Briefly, glass is coated with gold via a titanium intermediate, providing a plasmonic surface to enhance fluorescence signals emitted by TIRFM. The gold surface is reacted with thiol-poly(ethylene glycol)-biotin enabling the subsequent functionalization of NeutrAvidin (NA) and biotinylated antibodies and proteins targeting external epitopes of the single particles. Lastly, fluorescent-dye-conjugated antibodies were utilized to perform IF, and molecular beacons were utilized to perform FISH (**Fig. 1a**). TIRFM provides an evanescent wavefront that exponentially decreases from the coverslip surface, affording the visualization of biomolecules in single virions and IHD-EVs as localized fluorescent signals that can be quantified as functions of fluorescent emissions (**Fig. 1b**). As a model system, we chose SARS-CoV-2 for the validation of the BARA. Transmission electron microscopy (TEM) revealed the presence of spike glycoproteins forming the corona that is a hallmark of coronaviruses (**Fig. 1c**). Therefore, the spike glycoprotein along with targeting multiple regions of the nucleocapsid-encoding RNA via combining IF and FISH provided the colocalization of fluorescent signals on a single-localized domain, providing evidence for the co-expression of biomolecules on a single virion (**Fig. 1d**). While SARS-CoV-2 is the model system for validation, the tunable nature of the BARA is illustrated by simultaneous targeting antigens and nucleic acids for Influenza A (**Fig. 1e**) and the respiratory syncytial virus (RSV; **Fig. 1f**). Altogether, the BARA provides a tunable platform to multiplex antigenic and nucleic acid signals in single virions and IHD-EVs.

**Figure 1:**
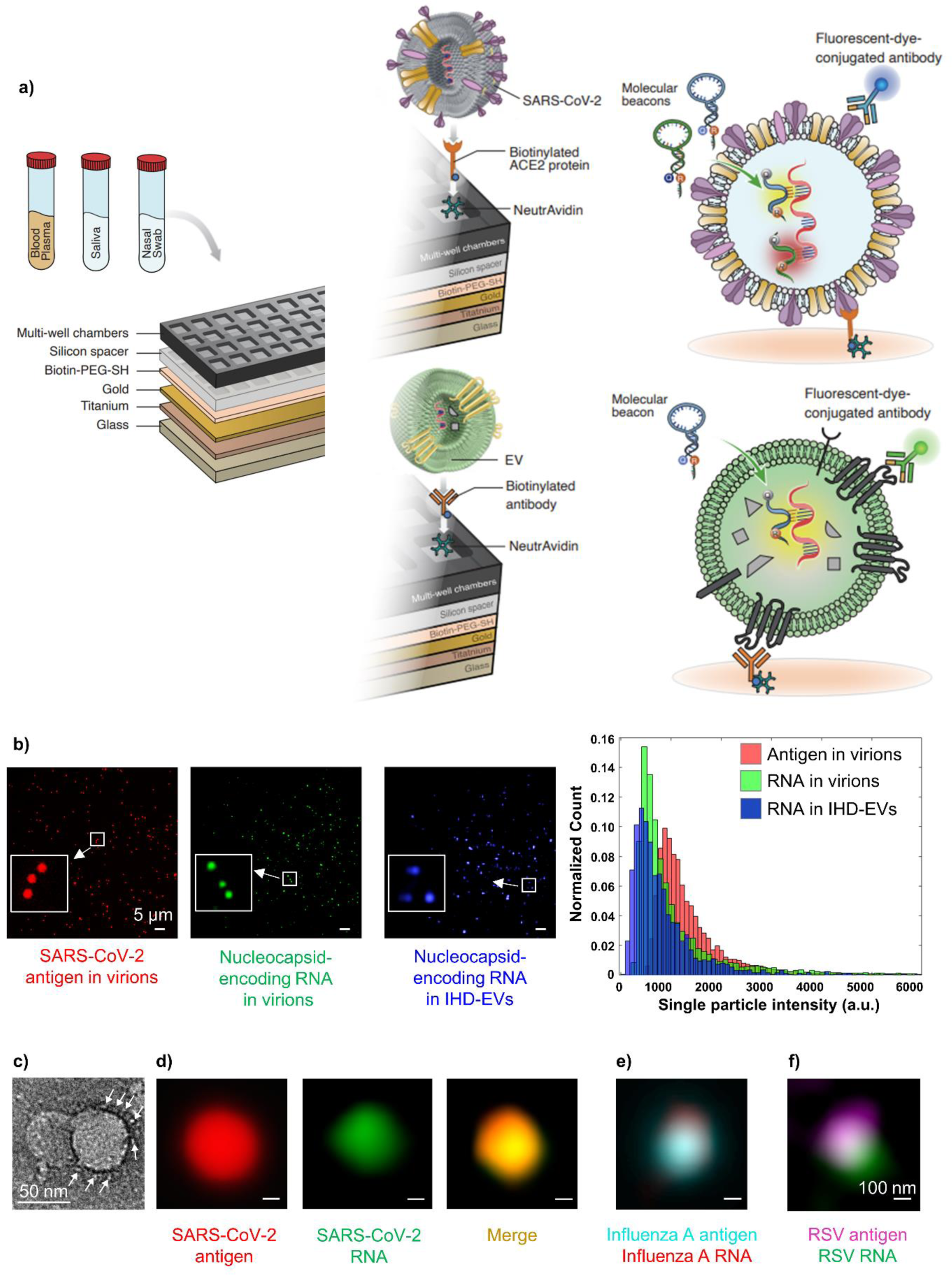
Single virion antigenic and nucleic acid detection with the BARA. **(a)** the BARA assembly and schematic mechanism for detecting antigens and nucleic acids utilizing immunofluorescence (IF) and fluorescent *in situ* hybridization (FISH), respectively. Detection is performed on single virions and infected extracellular vesicles (EVs) derived from complex biofluids, such as blood plasma, saliva, and nasopharyngeal (nasal) swabs (NS). **(b)** Representative total internal reflection fluorescence microscopy (TIRFM) images illustrating the spike glycoprotein on single virions and nucleocapsid-encoding RNA in single virions, and IHD-EVs, accompanied by their corresponding fluorescence intensity histograms. **(c)** Transmission electron microscope (TEM) image of SARS-CoV-2 virions showcasing the visible spike glycoprotein forming coronavirus corona. **(d)** Representative TIRFM images demonstrating SARS-CoV-2 antigen and nucleocapsid-encoding RNA on a single SARS-CoV-2 virion and the combination of detection methods, revealing colocalization of fluorescent signals in a localized domain. **(e)** Representative TIRFM images displaying the colocalization of antigens and nucleic acids for Influenza A. **(f)** Representative TIRFM images showing the colocalization of antigens and nucleic acids for respiratory syncytial virus (RSV).

### Specific and sensitive detection of single SARS-CoV-2 virions

To isolate single SARS-CoV-2 virions onto the plasmonic surface of the BARA, we tested different methods for immunopositive selection, including antibodies targeting the S1 and S2 subunits of the spike glycoprotein and the coronavirus membrane protein. On the other hand, immunopositive selection was also performed by simulating the port of entry for cellular infection (41, 42) with recombinant angiotensin-converting enzyme 2 (ACE2). Optimizing the surface for the capture of single virions, demonstrated that targeting the spike glycoprotein via antibodies or recombinant ACE2 provided the highest relative fluorescence intensity for IF of the spike glycoprotein (**Fig. S1a**, **Table S1**). Relative fluorescence intensity was defined as the sum of fluorescence intensity signals of the sample normalized by that of the negative control (phosphate-buffered saline, PBS) (37, 43). Scanning electron microscopy demonstrated the capture of single SARS-CoV-2 virions within the ACE2-functionalized surface of the BARA (**Fig. S1b**). Flow cytometry was performed on spike glycoprotein detection for SARS-CoV-2 virions and PBS to cross-validate the IF of the spike glycoprotein with the BARA, revealing a fluorescent enrichment for the sample only (**Fig. S2a**). For nucleic acid detection, specificity was tested with flow cytometry by targeting three regions of the nucleocapsid-encoding RNA on SARS-CoV-2, murine leukemia virus (MLV), and PBS. The molecular beacons hybridized at higher rates for the SARS-CoV-2 samples, whereas MLV and PBS yielded similarly low levels of fluorescent signal (**Fig. S2b**). Having detection methods for antigens and nucleic acids in SARS-CoV-2 virions, we tested the ability to colocalize signals by combining detection methods in single virions. Therefore, we utilized the BARA while co-targeting the spike glycoprotein and nucleocapsid-encoding RNA via *in situ* TIRFM image acquisition. Colocalized signals were observed that could be investigated multi-dimensionally. Apart from measuring fluorescence intensity one-dimensionally as total or relative fluorescence intensities, herein we demonstrate the spatial expression of the spike glycoprotein and nucleocapsid-encoding RNA on a single SARS-CoV-2 virion by measuring fluorescence intensity as a function of the *x – y* plane (**Fig. S3a**). Furthermore, cross-sections of the three-dimensional expression provide a two-dimensional fluorescence intensity profile as a function of an axis, revealing more spatially variable and diffuse expression of the spike glycoprotein compared to the nucleocapsid-encoding RNA (**Fig. S3a**).

Demonstrating an ability to distinguish antigenic and nucleic acid signals in single SARS-CoV-2 virions from negative controls, we aimed to characterize the sensitivities of the BARA. Therefore, we compared the BARA utilizing the top candidates for virion capture to the most sensitive COVID-19 diagnostic assay, qRT-PCR. Targeting S1 and S2 subunits of the spike glycoprotein exhibited a linear range of 10^3^–10^6^ particles/well (R^2^ = 0.99; ANOVA, *p* = 0.0023), whereas utilizing recombinant ACE2 to immobilize the single virions lent a linear range of 10^2^– 10^6^ particles/well (R^2^ = 0.98; ANOVA, *p* = 0.0008) when detecting the spike glycoprotein (**Fig. 2a**, **Fig. S3b**). Similarly, detecting nucleocapsid-encoding RNA when immobilizing with recombinant ACE2 demonstrated a linear range of 10^2^–10^6^ particles/well (R^2^ = 0.97; ANOVA, *p* = 0.0152). Therefore, both SARS-CoV-2 antigenic and nucleic acid detection via ACE2-mediated immobilization outperformed qRT-PCR by an order of magnitude, which became undetectable at 10^2^ particles/well (**Fig. 2a**). To further demonstrate the ability of the BARA to detect at the limit of detection (LoD), ACE2-mediated immobilization was coupled with both spike glycoprotein and nucleocapsid-encoding RNA to detect SARS-CoV-2 virions spiked into the healthy donor saliva at 10^2^ particles/well. Compared to healthy donor saliva, the single SARS-CoV-2 virions at the LoD demonstrated a significantly higher signal for spike glycoprotein detection (**Fig. 2b**; Welch’s two-tailed *t-*test, *p* < 0.0001). Furthermore, detection of the nucleocapsid-encoding RNA at the LoD further demonstrated significantly higher total fluorescence intensities than healthy saliva (**Fig. 2c**; Welch’s two-tailed *t-*test, *p* < 0.0001). Next, we tested whether freezing saliva affected the ability to detect the spike glycoprotein with the BARA. Therefore, SARS-CoV-2 virions were spiked at varying dilutions into saliva and tested immediately or frozen then rethawed. While there was an effect of freezing on the detection of various dilutions, such as a reduction in the slope (ANOVA, *p* < 0.0001 for the interaction effect), the BARA could discern the dilutions linearly for both frozen and fresh saliva samples (**Fig. 2c-d**; ANOVA, *p* = 0.0090 for frozen saliva and *p* = 0.0003 for fresh saliva). To further show the utility of the BARA as an automated high-throughput diagnostic assay, four BARA assays can be assembled in parallel alongside an automated pipetting machine for the facile testing of 256 samples. Various dilutions of SARS-CoV-2 virions were introduced, allowing for the testing of 256 samples at different LoD simultaneously (**Fig. 2e**). Utilizing the high-throughput technique, each sample corresponded with a total fluorescence intensity (**Fig. 2f**), which could be translated to SARS-CoV-2 positivity via higher total fluorescence intensities than that of the LoD (**Fig. 2g**). With this cutoff, the BARA yielded a positive percentage agreement (PPA) of 100 % and a negative percentage agreement (NPA) of 100 % (**Fig. 2g**).

**Figure 2:**
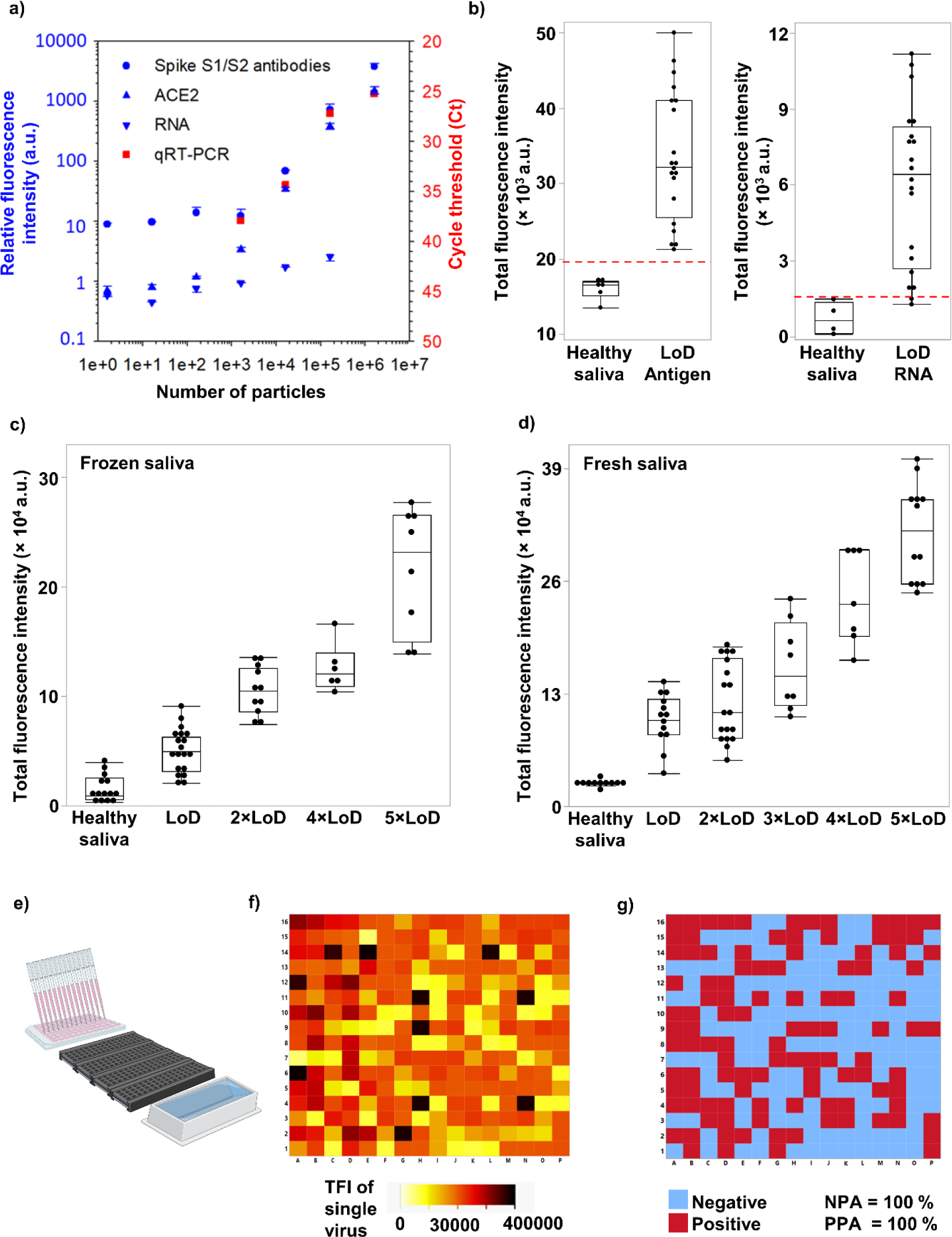
Performance evaluation of the BARA. **(a)** Comparative evaluation of antigenic and nucleic acid signals from the BARA in comparison to standard qRT-PCR. Inactivated SARS-CoV-2 virions were spiked in PBS at concentrations ranging from 0 to 1 × 10^6^ particles/well (n = 3, error bars indicate the standard deviation). **(b)** Detection results at the LoD of the BARA (n = 20 for LoD, n = 4 for pooled healthy donor saliva), demonstrating 20/20 detectable results for the antigen test and 19/20 detectable results for the RNA test. **(c)** Stability analysis of the BARA with frozen saliva specimens spiked at different LoDs (n = 69 with at least 10 replicates for 1-2 × LoD, 3-5 × LoD, and pooled healthy donor saliva conditions). **(d)** Stability analysis of the BARA with fresh saliva specimens spiked at different LoDs (n = 58 with at least 10 replicates for 1-2 × LoD, 3-5 × LoD, and pooled healthy donor saliva conditions). **(e)** Schematic representation of the high-throughput, automatic assay realized by the BARA, demonstrating the use of 4 coverslips in parallel on a plate holder and an automated pipetting machine for simultaneous testing of 256 samples. **(f)** Assessment of total fluorescence intensity of individual samples at various SARS-CoV-2 virion concentrations, ranging from 0 to 5 × LoD. **(g)** Interpretation of negative and positive samples after applying the cut-off value, resulting in a positive percentage agreement (PPA) of 100% and a negative percentage agreement (NPA) of 100%.

To prove specificity, we tested three times the LoD of SARS-CoV-2 virions for antigenic and nucleic acid detection with the BARA in the presence of multiple respiratory pathogens (**Table S2**). While the respiratory pathogens affected the fluorescent signal (ANOVA, *p* < 0.0001 for the interaction effect for both antigenic and nucleic acid detection), the BARA accurately discriminated between samples spiked with the SARS-CoV-2 virions and the corresponding control (**Fig. 3a**; ANOVA, *p* < 0.0001 for the effect of spiking for both antigenic and nucleic acid detection). Furthermore, three times the LoD in the presence of various endogenous and exogenous substances used for curing or lessening symptoms associated with SARS-CoV-2 infections was tested with the BARA (**Table S3**). Despite the endogenous and exogenous substance utilized affecting the fluorescence intensity (ANOVA, *p* < 0.0001 for the interaction effect for both antigenic and nucleic acid detection), the BARA distinguished the spiked samples from the control in the presence of endogenous and exogenous substances (**Fig. 3b**; ANOVA, *p* < 0.0001 for the effect of spiking for both antigenic and nucleic acid detection). Lastly, to test the ability to disseminate the BARA, accelerated stability was performed according to the Clinical and Laboratory Standards Institute (CLSI) EP25-A to evaluate the stability of *in vitro* diagnostic reagents (**Table S4-5**). The entire assay, including the biochip and the reagents therein, were incubated and tested at various temperatures and time points to determine the efficiency of detecting the spike glycoprotein on single SARS-CoV-2 virions (**Fig. S4a**). Utilizing the Arrhenius equation, various rate constants for each temperature were extracted from which the rate of degradation at 4 °C was extrapolated (**Fig. S4b-c**), revealing a 10 % degradation of the BARA at 94.26 days when stored at 4 °C. Therefore, the BARA holds diagnostic promise as an automated high-throughput clinically relevant assay for simultaneously detecting antigens and nucleic acids in single SARS-CoV-2 virions with sensitivities higher than qRT-PCR.

**Figure 3:**
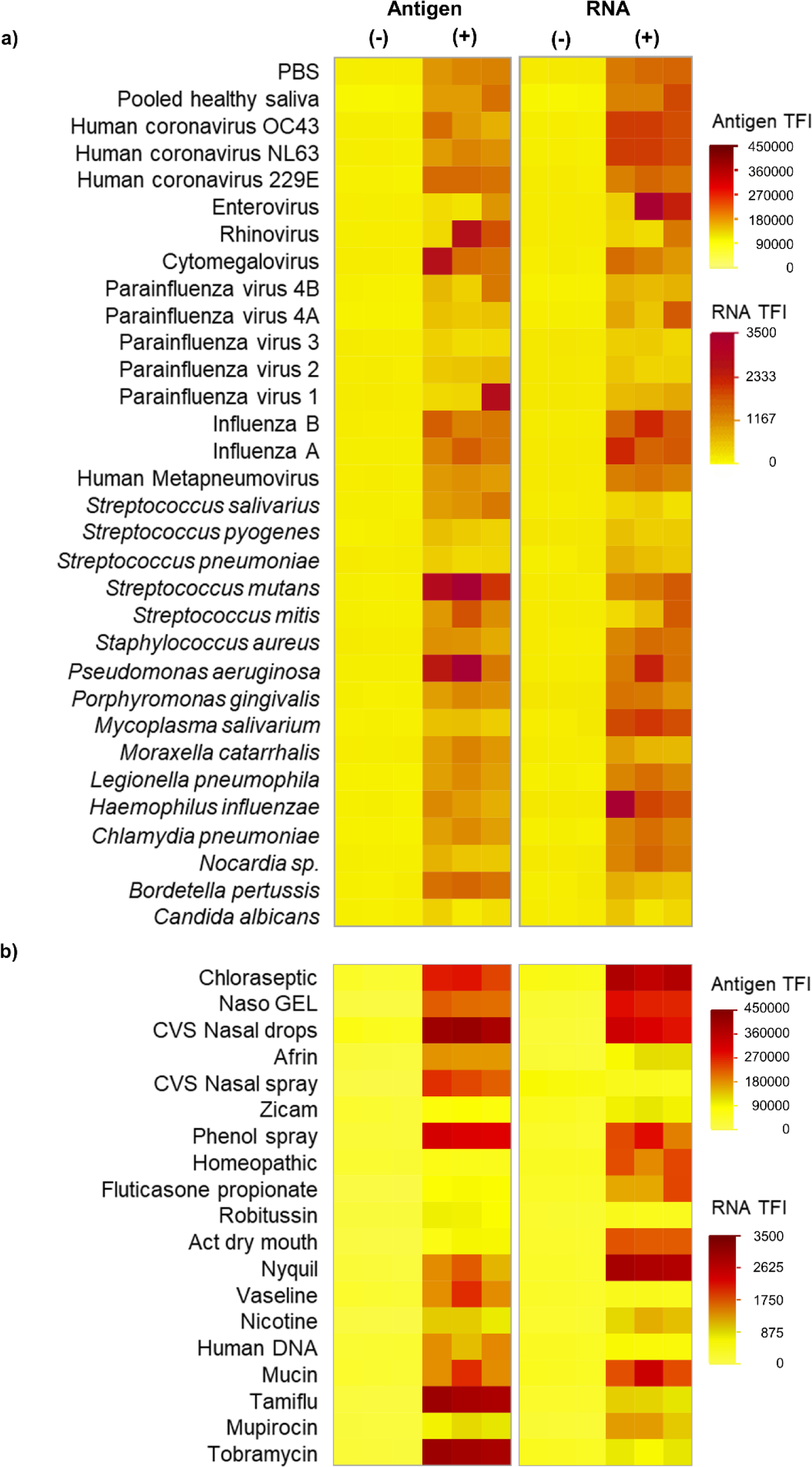
Cross-reactivity, microbial interference, and endogenous/exogenous substances interference. **(a)** Heatmap of antigen and nucleocapsid-encoding RNA signals during the evaluation of various respiratory pathogens, including 14 viruses, 15 bacteria, and 1 fungus at various concentrations **(Table S2)**, in the presence of 0 and 3 × LoD of SARS-CoV-2 virions with antigenic and nucleic acid detection for three replicates. **(b)** Heatmap of spike glycoprotein and nucleocapsid-encoding RNA signals during the evaluation of various endogenous/exogenous substances, including 16 substances and 3 common antiviral and antibacterial medicines, spiked into healthy saliva samples at different concentrations **(Table S3)**, in the presence of 0 and 3 × LoD of SARS-CoV-2 virions with antigenic and nucleic acid detection for three replicates.

### Monitoring genetic mutations on single virions

Given that SARS-CoV-2 variants alter the structure of the spike glycoprotein to evade immune responses (44), we tested the ability of the BARA to detect the spike glycoprotein on the various variants experienced during the pandemic in the USA, including the original Washington strain (USA-WA1/2020), alpha, beta, gamma, delta, and omicron strains. Despite the mutations, positive signals for the spike glycoprotein were obtained for all strains (**Fig. 4a**). Furthermore, the BARA could distinguish positive signals for all strains from the negative control of PBS (**Fig. S5a**; Dunnett’s test, *p* ≤ 0.0162). Since variants arise from mutations in the viral genome, we wanted to test whether the enhanced sensitivity of the BARA would be sufficient to detect genetic mutations at a single-virion resolution. Therefore, we designed molecular beacons to co-target S2-encoding RNA and variant-specific mutations for delta and omicron variants emitting different wavelengths upon TIRFM excitation. First, we determined whether the BARA could discern the ΔF157 and L452R mutations present in the delta variant from the Washington strain. The ΔF157 mutations revealed signal enrichment for the delta variant as opposed to the original Washington strain (Tukey’s HSD, *p* = 0.0045). However, the Washington strain emitted a higher total fluorescence intensity than the control (**Fig. S5a**; Tukey’s HSD, *p* = 0.0018). On the other hand, the L452R mutation was overexpressed in the delta strain as opposed to the original Washington strain (Tukey’s HSD, *p* = 0.0106) and was absent for the Washington strain (**Fig. S5a**; Tukey’s HSD, *p* = 0.3309). Therefore, for the delta variant, the BARA colocalized signals for the spike glycoprotein, S2-encoding RNA, and the L452R mutation whereby the colocalization of fluorescent signals as the primary colors of light added to white light, demonstrating the co-expression of the biomolecules on a single virion (**Fig. 4b**). Similarly, the BARA colocalized signals for the spike glycoprotein, S2-encoding RNA, and the ΔH69 mutation on single virions (**Fig. 4b**), revealing an enrichment in the omicron variant when compared to the original Washington strain (**Fig. S5b**; Tukey’s HSD, *p* = 0.0018), albeit detectable in the original Washington strain (Tukey’s HSD, *p* = 0.0085). While the BARA distinguished positive signals for the different variants, to further determine the robustness of spike glycoprotein detection, multiple Pango lineages of the omicron variant were tested, including BA.5.1, BF.7, BQ.1, XBB.1.5, BA.2.12.1, and BA.2.3. Positive signals were observed for the Pango lineages (**Fig. 4c**) and were significantly higher than the negative control for BA.2.12.1, BA.5.1, and XBB.1.5 (**Fig. S5b**; Dunnett’s test, *p* ≤ 0.0487).

**Figure 4:**
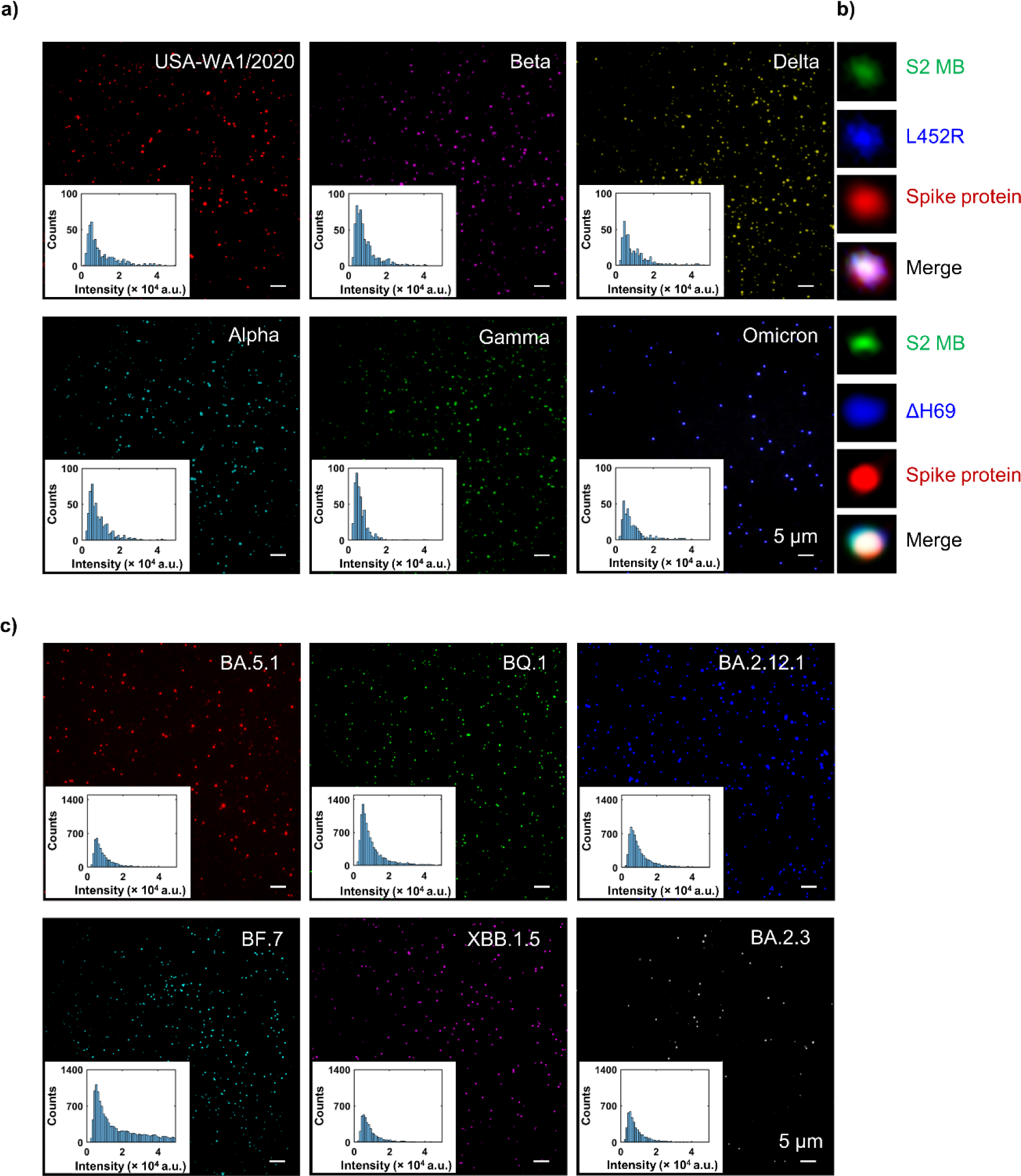
Lineage and mutation monitoring with the BARA. **(a)** Representative images of various SARS-CoV-2 variants, including the original Washington strain (USA-WA1/2020), alpha (B.1.1.7), beta (B.1.351), gamma (P.1), delta (B.1.617.2), omicron (BA.1) variants. Inset histograms illustrate the expression distributions of single virions as distributions of fluorescence intensity. **(b)** Representative image of colocalized signals for spike glycoprotein, S2-encoding RNA, and the L452R mutation in a single virion of the delta variant. Additionally, colocalized signals for the spike glycoprotein, S2-encoding RNA, and the ΔH69 single point mutation in a virion of the omicron variant. **(c)** Variant monitoring by detecting various Pango lineages of the omicron variant using the BARA for spike glycoprotein detection, including BA.5.1, BF.7, BQ.1, XBB.1.5, BA.2.12.1, and BA.2.3. Inset histograms illustrate the expression distributions of single virions as distributions of fluorescence intensity.

### Dual antigenic and nucleic acid detection for COVID-19 diagnosis

To test the clinical potential of the BARA for virion-mediated infections, we validated its use for distinguishing COVID-19 patients utilizing complex biofluid samples, including saliva (n = 33 patients, n = 30 healthy donors) and NS (n = 40 patients, n = 19 healthy donors). Based on clinical testing, patient specimens were collected from PCR-confirmed COVID-19 cases (**Table S6-7**). Spike glycoprotein and nucleocapsid-encoding RNA were utilized to determine COVID-19-positive patients. Both antigenic and nucleic acid detection on single SARS-CoV-2 virions demonstrated an enhanced signal for COVID-19 patients for saliva (**Fig. 5a**; Mann-Whitney *U* test, *p* < 0.0001 for antigenic and nucleic acid detection) and NS (**Fig. 5a**; Mann-Whitney *U* test, *p* < 0.0001 for antigenic detection, *p* = 0.0002 for nucleic acid detection). Specifically, spike glycoprotein detection in salivary single SARS-CoV-2 virions revealed a sensitivity of 94 % and a specificity of 100 %, whereas nucleocapsid-encoding RNA detection yielded a sensitivity of 91 % and a specificity of 100 % (**Fig. S6a**). On the other hand, detection of nasopharyngeal single SARS-CoV-2 virions provided sensitivities of 88 % and 80 % for spike glycoprotein and nucleocapsid-encoding RNA detection, respectively, and specificities of 100 % for both detection methods (**Fig. S6b**). Given that the highest specificity was obtained for spike glycoprotein detection in salivary single SARS-CoV-2 virions, we wanted to see whether the enhanced sensitivity of the single-virion method could diagnose the COVID-19 asymptomatic patient subpopulation (45, 46). Therefore, a cohort of 20 asymptomatic patients was tested with the BARA, revealing an enrichment of signals for the asymptomatic cohort compared to healthy donors (**Fig. 5b**; Mann-Whitney *U* test, *p* = 0.0003).

**Figure 5:**
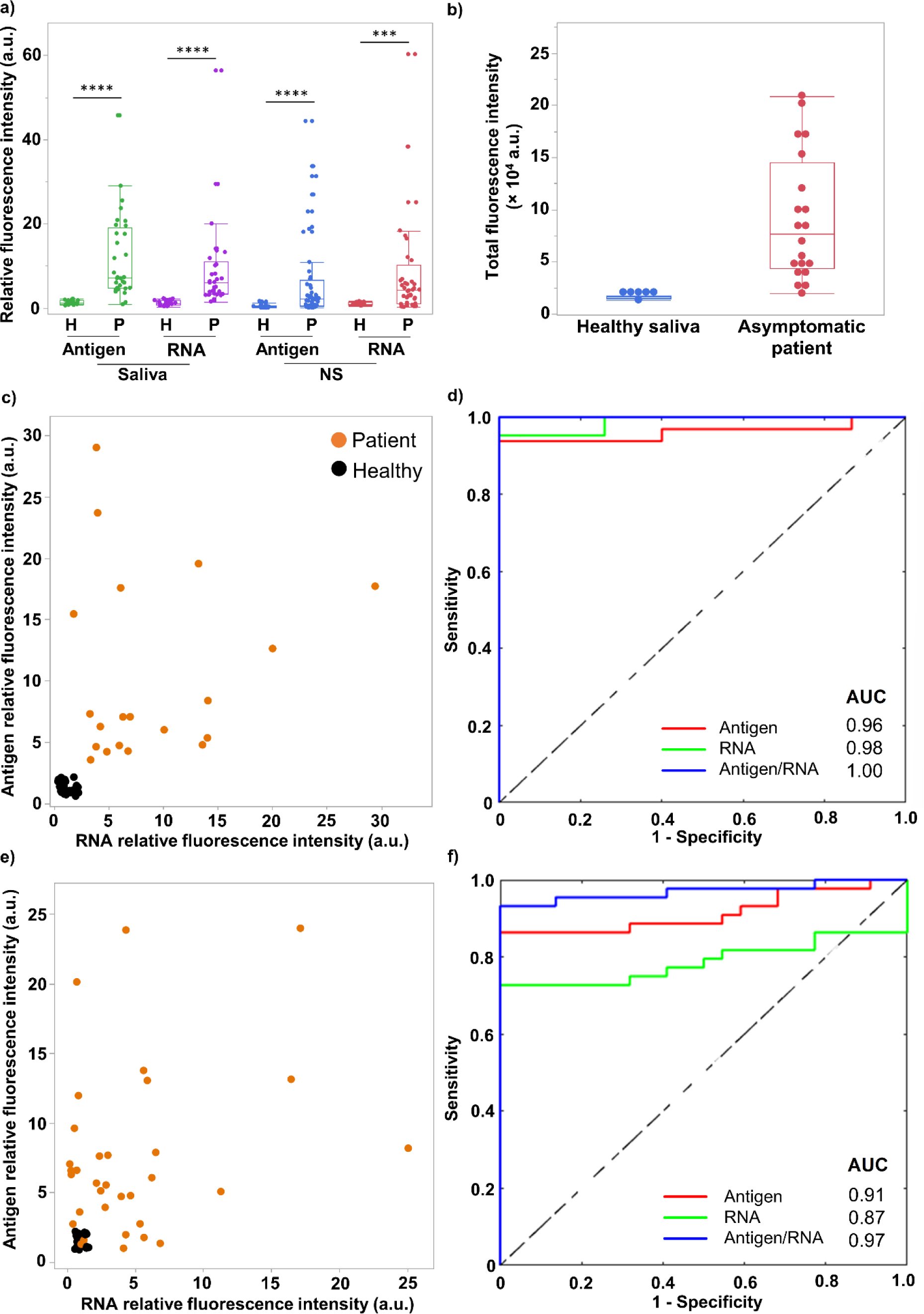
Clinical validation of the BARA using saliva and NS specimens from COVID-19-positive patients. **(a)** Antigenic and nucleic acid signals from COVID-19-positive patients are significantly enhanced in comparison to healthy donor (Mann-Whitney *U* test, \*\*\*\**p* <0.0001, \*\*\**p* <0.001) in both saliva (n = 33 patients, n = 30 healthy donors) and NS specimens (n = 40 patients, n = 19 healthy donors). **(b)** Clinical validation of asymptomatic COVID-19-positive patient saliva (n = 20 for asymptomatic, n = 6 pooled healthy donor saliva). **(c)** Correlation between antigenic and nucleic acid expression of saliva specimens, with minimal expression observed in the saliva of healthy donors (Pearson’s correlation coefficient, *p* < 0.0001 for *r* = 0.76). **(d)** Receiver operating characteristic (ROC) curves for combined detection of antigens and nucleic acids in salivary single SARS-CoV-2 virions with an enhanced area under the curve (AUC) value of 1.00 for dual detection. (**e)** Correlation analysis of antigenic and nucleic acid expression of nasopharyngeal single SARS-CoV-2 virions, revealing a subpopulation of healthy donors with minimal expression of both biomarkers. **(f)** ROC curve analysis of antigenic, nucleic acid, and the dual biomarker expression in NS specimens, indicating an enhanced AUC value of 0.97.

To further improve the sensitivities of the BARA as NS demonstrated lower diagnostic performances, we utilized the ability of the BARA to dual detect antigenic and nucleic acid signals to provide a combinatorial approach to diagnose COVID-19 for salivary and nasopharyngeal samples. Antigenic and nucleic acid expression strongly coincided (Pearson’s correlation coefficient, *p* < 0.0001 for *r* = 0.76) and had weak negative associations with qRT-PCR cycle-threshold values for COVID-19 patients (Pearson’s correlation coefficient, *r =* −0.33 for antigenic detection, *r =* −0.14 for nucleic acid detection), revealing a distinct subpopulation for healthy donors with minimal expression of both biomarkers (**Fig. 5c**). Utilizing receiver operating characteristic (ROC) curves, the combined detection of antigens and nucleic acids in salivary single SARS-CoV-2 virions augmented the area under the curve (AUC) from 0.96 for antigenic detection and 0.98 for nucleic acid detection to 1.00 for dual detection (**Fig. 5d**). Thus, the sensitivity was enhanced to 100 % (**Fig. S6b**). Despite the expression of nasopharyngeal single SARS-CoV-2 virions only weakly associating between antigenic and nucleic acid detection (Pearson’s correlation coefficient, *r =* 0.15) with weak negative correlations with qRT-PCR cycle-threshold values for COVID-19 patients (Pearson’s correlation coefficient, *r =* −0.29 for antigenic detection, *r =* −0.44 for nucleic acid detection), a subpopulation of healthy donors with minimal expression of both biomarkers was present with slight intercalations of COVID-19 patients (**Fig. 5e**). ROC curves revealed the enhanced diagnostic capability of the BARA by utilizing the combinatorial method, increasing the AUC from 0.91 and 0.87 for single antigenic and nucleic acid detection, respectively, to 0.97 for dual detection (**Fig. 5f**), which increased the sensitivity to 95 % (**Fig. S6b**). Therefore, dual antigenic and nucleic acid detection with the BARA enhances sensitivities for a reliable COVID-19 diagnostic assay.

### Virion-RNA detection in plasma-derived EVs of post-acute sequelae of COVID-19 (PASC) patients

Patients with COVID-19 may suffer a heterogeneous set of symptoms post-infection, which is referred to as PASC (47), ranging from neurologic (48) to cardiovascular symptoms (49), and affecting various organs (50). Therefore, we hypothesized that virion-RNA may be present in EVs of PASC patients originating from infected tissue. PASC patients were recruited as patients with ongoing, relapsing, or new symptoms persisting beyond 30 days of acute infection. Samples from seven patients with PASC were used in the investigation (**Table S8-9**), whereby their plasma was collected serially at three time points and co-detected with qRT-PCR and the BARA. Tunable resistive pulse sensing (TRPS) on the size distribution of EVs from saliva and plasma revealed similar profiles to SARS-CoV-2 virions (**Fig. S7**). Therefore, it was necessary to specifically isolate EVs from SARS-CoV-2 virions. The use of an antibody cocktail targeting CD63 and CD9, which are tetraspanins enriched in various subpopulations of EVs (51), revealed an absence of signal in a patient saliva sample with COVID-19, but an enrichment of CD63^+^ single EVs (**Fig. S8**). On the other hand, capturing particles with antibodies targeting the S1 and S2 subunits of the spike glycoprotein revealed a loss of CD63^+^ single EVs (**Fig. S8**). Furthermore, to retrieve EVs from plasma, we conducted various isolation methods, including dextran-based precipitation, size-exclusion chromatography, and thrombin for the cleavage of fibrinogen (52). The EVs isolated with thrombin produced CD63 and CD81 signals at higher frequencies compared to the other isolation methods (**Fig. S9a-b**), which had less loss of EVs than the other protocols (**Fig. S9c**).

To test the presence of virion-RNA in IHD-EVs, thrombin-treated plasma from PASC patients was screened with the BARA whereby EVs were isolated via positive immunoselection with CD63/CD9-targeting antibodies and detected for the nucleocapsid-encoding RNA. A high-intensity subpopulation for nucleocapsid-encoding RNA expression was detected in single IHD-EVs after long timeframes extending past 200 days, which was absent in healthy donor serum, while CD63 expression remained the same in single EVs across healthy donor and PASC patient samples (**Fig. 6a**). Total expression of CD63 and the nucleocapsid-encoding RNA demonstrated that the detection of IHD-EVs was EV-independent, as CD63 signals remained high for healthy donors and PASC patients (Student’s two-tailed *t-*test, *p* = 0.35), whereas nucleocapsid-encoding RNA signals were higher for PASC patients (**Fig. 6b**; Student’s two-tailed *t-*test, *p* = 0.0055). Furthermore, the presence of CD63^+^ IHD-EVs containing nucleocapsid-encoding RNA was demonstrated by their colocalization on single IHD-EVs (**Fig. 6c**). Interestingly, colocalization analysis with the BARA revealed that the loading percentage of nucleocapsid-encoding RNA in IHD-EVs decreased over the course of infection (**Fig. 6e**). Although qRT-PCR and the BARA did not correlate in levels of expression for the nucleocapsid-encoding RNA in IHD-EVs (Pearson’s correlation coefficient, *r =* 0.08), likely due to qRT-PCR being at the cusp of its LoD (**Table S8**), the qRT-PCR demonstrated positivity whereby the BARA provided more holistic profiles of the nucleocapsid-encoding RNA levels as a function of time (**Fig. 6d**). Therefore, the BARA demonstrated the packaging of virion-RNA in IHD-EVs of patients with symptoms post-infection, possibly describing residual virion-RNA as the culprit for the long-term symptoms associated with PASC (50, 53).

**Figure 6:**
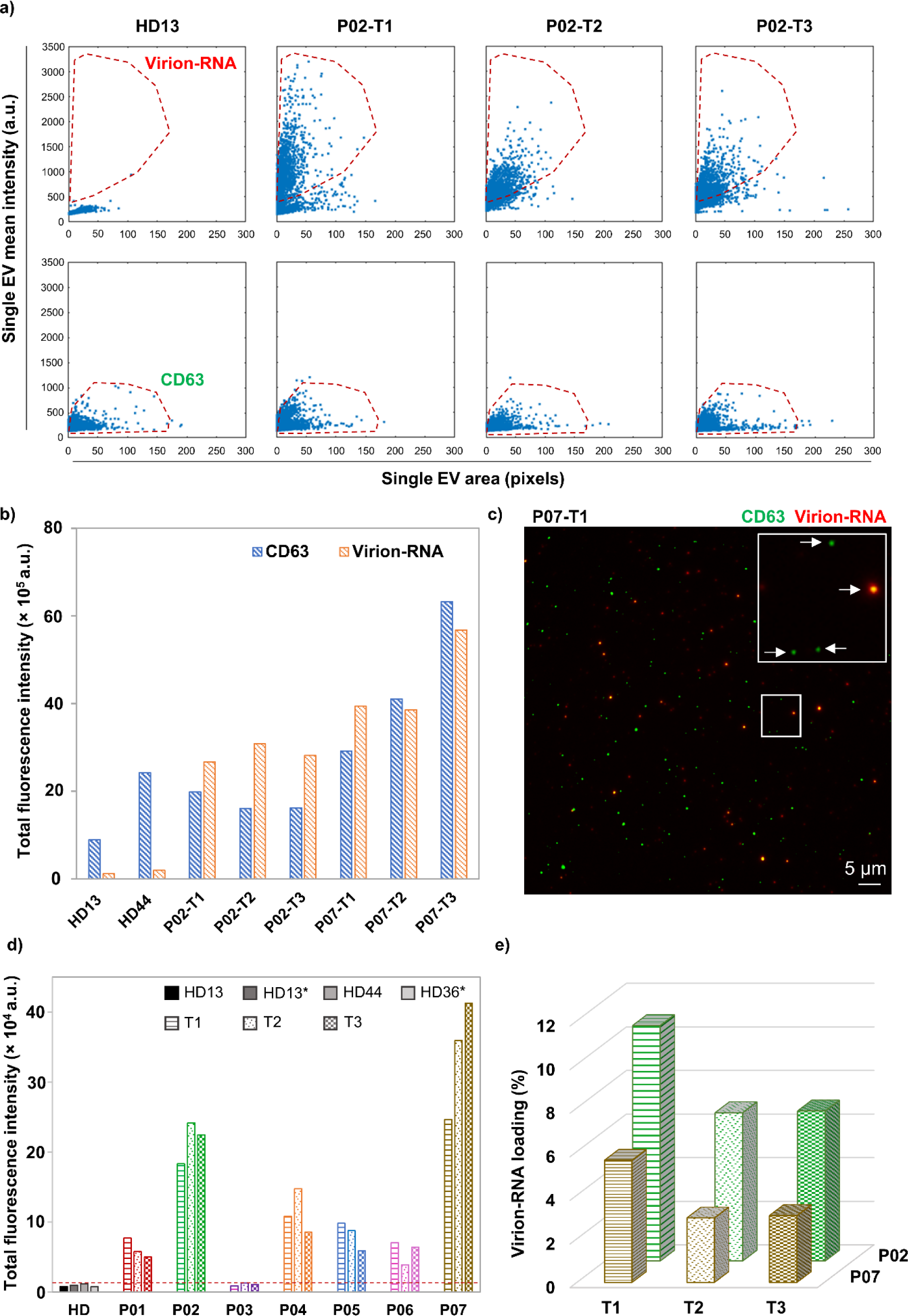
Virion-RNA detection in IHD-EVs of PASC patients. **(a)** Comparison of virion-RNA content and the CD63 expression in single IHD-EVs compared to healthy donor plasma. **(b)** Dual detection of CD63 and nucleocapsid-encoding RNA in single EVs of healthy donors and PASC patients P02 and P07. **(c)** Colocalization analysis of CD63 and nucleocapsid-encoding RNA signals in single IHD-EVs. Inset demonstrates CD63^+^ IHD-EVs containing nucleocapsid-encoding RNA as highlighted by the white arrows. **(d)** Detection of nucleocapsid-encoding RNA in thrombin-treated plasma of seven PASC patients (P01 to P07) at 3 serial timelines (T1, T2, and T3). A comparison with thrombin-treated plasma from donors who fully recovered from COVID-19 (HD13 and HD13*, indicating 1-month duration, HD44) and thrombin-treated plasma from a healthy donor (HD36, collected in 2019) prior to the COVID-19 outbreak. **(e)** Virion-RNA loading percentage in CD63^+^ IHD-EVs at different stages of infection.

## Discussions

The BARA demonstrates the translation of single-EV technologies for clinical application. Progressing toward single-EV detection has afforded unprecedented sensitivities surpassing enzyme-linked immunosorbent assay (ELISA) for protein detection (37) and qRT-PCR for microRNA (43) and messenger RNA detection (37). Herein, the BARA outperformed qRT-PCR by detecting the spike glycoprotein and nucleocapsid-encoding RNA in single SARS-CoV-2 virions. The enhanced sensitivity allowed for the detection of a subpopulation of COVID-19 patients that often eluded qRT-PCR tests, which were asymptomatic and were partly responsible for the vast undetectable spread of the SARS-CoV-2 virion (54–56). Another subpopulation of COVID-19 patients that were partly responsible for the spread of the disease was recently infected individuals who often presented false-negative results (57–59). Detecting patients upon infection before replication whereby virion levels are too low for contagion is a diagnostic window to limit the spread of virion-mediated outbreaks by enforcing a bottleneck to virion transmission, such as is observed for highly mortal pathogenic diseases like Ebola (60). Therefore, single-virion methods may aid in the rapid detection and subsequent mitigation of outbreaks via efficiently detecting patients with low-virion counts.

Another benefit to utilizing single-EV methods is the facile deconvolution of vesicular heterogeneity via *in situ* colocalization of biomolecular signals (33, 61). The BARA offered a unique qualitative perspective through the colocalization of biomolecular signals, allowing the visualization of mutation-harboring virions and virion-RNA within IHD-EVs in PASC patients. Due to the growing risk of zoonoses, an active field in emerging infective diseases is the identification of mutations that may lead to animal-to-human transmission or *vice versa* (62). The BARA can aid in identifying rates of mutation and the likelihood of zoonoses via the serial colocalization of human-infecting genes with a housekeeping gene in various animals that pose a threat to interspecies spillover. On the other hand, with the BARA we determined that PASC patients contain virion-RNA in IHD-EVs after extended timeframes. Given the heterogeneous symptoms experienced by PASC patients (63–65), colocalizing organ-specific biomarkers with EV biomarkers and virion-RNA may elucidate the causations of symptoms or even predict symptoms before their onset. Moreover, antibodies can be screened in high throughput via IF on various virion strains to determine the efficacy of vaccine targets. The avenues for the BARA open various investigative opportunities, many of which are currently being explored.

Lastly, the tunability of the BARA provides a customizable framework for many virion-mediated diseases. Although the investigation focused on detecting single SARS-CoV-2 virions to demonstrate the diagnostic capabilities of the BARA, selecting antibody cocktails corresponding to external epitopes of virions can be rapidly achieved. Herein, we demonstrated the ability of the BARA to detect antigens and nucleic acids simultaneously in single Influenza A and RSV virions by further designing molecular beacons and fluorescent-dye-conjugated antibodies tailored to the respective biomolecules. Apart from providing the unique multifaceted detection of single virions, the combination of IF and FISH for detecting antigens and nucleic acids in single virions enhanced the sensitivity of the BARA compared to single biomolecule detection. The increased sensitivities afforded the detection of single SARS-CoV-2 virions in complex biofluids and the sensitive identification of COVID-19 patients insofar as sensitivities of 100 % in the case of saliva samples were observed. The BARA provides a highly sensitive diagnostic assay that can be easily tuned to future outbreaks and provides a perspective into the pathophysiology of virion-mediated diseases, realizing a comprehensive assay unmet by current diagnostics.

## Materials and Methods

### The BARA fabrication

High-precision, 24 × 75 × 0.15 mm, borosilicate glass coverslips (D 263® M; Schott AG, Mainz, Germany) were cleaned with ethanol followed by deionized (DI) water in an ultrasonic bath for 5 min each. After repeating the cleaning process, the coverslips were dried with nitrogen gas. The coverslips were cleaned with a UV-ozone cleaner (Jelight, Irvine, CA) for 15 min. A 2-nm film of titanium was first deposited onto the cleaned coverslips via electron beam evaporation (DV-502A; Denton Vacuum, Moorestown, NJ) to facilitate the adhesion of gold to the surface. Utilizing the same technique, a 10-nm film of gold was deposited atop the titanium layer. The gold-coated coverslips were then submerged into a solution of thiolated molecules to functionalize the gold with biotin motifs, which was comprised of β-mercaptoethanol (βME; Sigma-Aldrich, St. Louis, MO), 2 kDa methoxy-poly(ethylene glycol)-thiol (mPEG-SH; Laysan Bio, Arab, AL), and 2 kDa biotin-PEG-SH (Nanocs, New York, NY) at a molar ratio of 95:3:2, respectively, in 200 proof ethanol (Thermo Fisher Scientific, Waltham, MA). The coverslips were incubated in the solution overnight at room temperature in a dark environment. Excess thiolated molecules were rinsed away with ethanol. The biotin-functionalized, gold-coated coverslips (referred to as biochips) were then dried with nitrogen gas and fastened to a 64-well ProPlate® microarray system (Sigma-Aldrich, St. Louis, MO).

### Antibody functionalization of the biochip surface

The working volume utilized for an individual well was 20 μL, which was kept constant for the different solutions added into the wells. Furthermore, all incubation steps were performed on a shaker to ensure a uniform coating of the solution throughout the well surface. Before antibody functionalization, each well was washed by pipetting DI water up and down 10 times. Then, a 50-μg/mL solution of NeutrAvidin (NA; Thermo Fisher Scientific) diluted in PBS (Thermo Fisher Scientific) was added into each well and incubated at room temperature for 1 hr to bind to the biotin motifs functionalized on the gold surface of the biochip. Excess NA was rinsed away by pipetting PBS up and down 10 times. The rinsing process was repeated three times. Capture antibodies and recombinant proteins previously biotinylated via the EZ-Link micro Sulfo-NHS-biotinylation kit (Thermo Fisher Scientific) were diluted to a concentration of 10 μg/mL in a 1 % (w/v) solution of bovine serum albumin (BSA; Sigma-Aldrich) in PBS. Specific capture antibodies or recombinant proteins to immobilize subpopulations (**Table S1**) were added into each well and incubated at room temperature for 1 hr. Excess antibodies and recombinant proteins were rinsed away by pipetting PBS up and down 10 times for three repetitions.

### Saliva collection

Following Institutional Review Board protocol 2021H0246 (Biomedical Sciences Committee at The Ohio State University), a de-identified cohort comprising 30 healthy donors, 33 symptomatic patients, and 20 asymptomatic patients was enrolled from which saliva samples were collected. Pooled healthy donor saliva was defined as the combined saliva from five healthy donors. After collection, the saliva was centrifuged at 2000 × *g* for 10 min and stored at −80 °C. All saliva samples were inactivated at 56 °C for 30 min prior to purification.

### Nasopharyngeal swab (NS) collection

A de-identified cohort of 19 healthy donors and 40 patients was collected following the Institutional Review Board protocol 2021H0246 (Biomedical Sciences Committee at The Ohio State University). All NS samples were inactivated at 56 °C for 30 min prior to purification.

### Plasma collection

Plasma samples were provided from a cohort of 7 COVID-19 patient participants with written informed consent, in accordance with the Code of Federal Regulations Title 45: Public Welfare Part 46: Protection of Human Subjects (45 CFR 46) (66). After collection, the plasma samples were stored at −80 °C. All plasma samples were inactivated at 56 °C for 30 min prior to purification. Defibrination of plasma samples was performed by adding 4.4 U/mL of thrombin (TMEXO-1, System Biosciences) to the plasma samples, incubating for 5 min, centrifuging at 10000 × *g* at room temperature for 5 min, and collecting the supernatant.

### Microorganism collection

Viruses, bacteria, and fungi were obtained from the Biodefense and Emerging Infections (BEI) Resources Repository, American Type Culture Collection (ATCC), and the Department of Pathology at The Ohio State University Wexner Medical Center. All viruses, bacteria, and fungi were inactivated at 56 °C for 30 min and diluted to the tested concentrations (**Table S2**).

### Biofluid sample purification

Biofluid samples were purified via size-exclusion chromatography (qEV; Izon Science, Christchurch, New Zealand) according to the manufacturer’s instructions. Briefly, a 200 µL biofluid sample was introduced through the column pre-wetted with PBS, whereby fractions 7 – 12 were collected. The purified samples were then re-concentrated to 200 µL with spin columns (10 kDa MWCO, Millipore Sigma Amicon Ultra Centrifugal Filter Unit, Fisher Scientific) at 4 °C at 3000 × *g*.

### Molecular beacon hybridization to membrane-enveloped virion RNA

Molecular beacons (**Table S10**) were diluted to a concentration of 5 μM in 12.5 × Tris EDTA (TE) buffer (Sigma-Aldrich) diluted in DI water to stabilize the molecular beacons and permeabilize the membrane encasing the RNAs (43). The molecular beacon cocktail was diluted 25 times within the purified biofluid sample and allowed to incubate for 2 hr at 37 °C in a dark environment to facilitate molecular beacon hybridization to the target RNA.

### Capture of virions and extracellular vesicles (EVs)

A 3 % (w/v) solution of BSA in PBS was incubated in each well at room temperature for 1 hr to block non-specific particle capture. After the removal of BSA, the biofluid samples containing virions or EVs (including pre-hybridized and untreated samples) were subsequently incubated in the wells of the BARA for 2 hr at room temperature in a dark environment. For the untreated samples, excess virions and EVs were washed by pipetting PBS up and down 10 times for a total of 3 repetitions. For the pre-hybridized samples, PBS was incubated in the wells for 5 min then pipetted up and down 10 times to remove excess virions and EVs and unhybridized molecular beacons. The rinsing step was repeated 4 times in a dark environment.

### Immunofluorescence of membrane proteins

A 3 % (w/v) solution of BSA in PBS was incubated in each well at room temperature for 1 hr to block the non-specific adhesion of the fluorescent-dye-conjugated antibodies. After withdrawing the BSA solution, a 1-µg/mL solution of the fluorescent-dye-conjugated antibodies (**Table S1**) in 10 % (w/v) normal goat serum (NGS; Thermo Fisher Scientific) was subsequently incubated in the wells for 1 hr at room temperature in a dark environment. Excess fluorescent-dye-conjugated antibodies were rinsed and removed by incubating in PBS for 5 min then pipetting up and down the solution 10 times. The rinsing step was repeated 3 times in a dark environment.

### Image acquisition and processing

A 10 × 10 array of images was acquired via TIRFM (Nikon, Melville, NY) for each well with a 100 × objective and immersion oil to reduce surface refraction. Exposure times and laser power were maintained across experiments to ensure the consistency of the assay. TIRFM images were quantified by measuring the total and mean fluorescence intensity of each spot detected by TIRFM. Histograms were generated with the total and mean intensities of the single spots detected by TIRFM. Scatter plots were generated with the mean intensity and size of the single spots detected by TIRFM. Relative and total fluorescent intensities of the sample were obtained from custom-built algorithms that were previously reported (31, 37).

### Real-time quantitative reverse transcription-polymerase chain reaction (qRT-PCR)

Membrane-enveloped RNA was isolated with the miRNeasy Serum/Plasma kit (Qiagen, Hilden, Germany) and the Single Cell RNA Purification Kit (Sigma-Aldrich) according to the manufacturer’s instructions. The isolated RNA was combined with random primers (Thermo Fisher Scientific) and was heated to 70 °C for 2 min to ensure that the target RNA was single-stranded and cooled to 4 °C to anneal the primers. A solution containing Moloney murine leukemia virus reverse transcriptase (MMLV-RT; Thermo Fisher Scientific) and deoxyribonucleotide triphosphate (dNTP; Thermo Fisher Scientific) in a buffer comprised of dithiothreitol (DTT; Thermo Fisher Scientific), RNaseOUT™ Recombinant Ribonuclease Inhibitor (Thermo Fisher Scientific), and Maxima First Strand cDNA Synthesis Kit Reaction Mix (Thermo Fisher Scientific) was heated to 42 °C for 1 hr to synthesize cDNA, then heated to 95 °C for 5 min to deactivate the RT, and subsequently cooled to 4 °C for storage purposes. The cDNA was introduced to a TaqMan™ Fast Advanced Master Mix (Thermo Fisher Scientific) and combined with probes targeting the open reading frame (ORF1ab), the spike protein, the nucleocapsid protein, and the human ribonuclease P RNA component H1 gene (RPPH1; RNase P) as a positive control as provided by the TaqMan™ 2019-nCoV Assay Kit v1 (Thermo Fisher Scientific). Real-time qRT-PCR was performed on the sample with an activation step of 95 °C for 20 s followed by 45 cycles of denaturing at 95 °C for 3 s and annealing and extending at 60 °C for 30 s.

### Transmission electron microscopy (TEM)

Two 20 µL droplets of water for injection (WFI) and two 20 µL droplets of negative stain (UranyLess EM stain, Electron Microscopy Sciences) were placed on a strip of parafilm. The TEM grid was subjected to plasma treatment for 1 min. Then, 10 µL of a SARS-CoV-2 virion solution was carefully placed onto the treated surface of the grid. The virions solution was incubated on the surface of the grid surface for 1 min and gently blotted with filter paper to remove excess liquid. The virion-coated grids were submerged into the WFI droplet and blotted dry using filter paper. The process was repeated with the second WFI droplet. The virion-coated grids were stained via submersion into the first droplet of negative stain, followed by blotting, and then submerging again into the second droplet of the negative stain. The grid was allowed to incubate in the stain for approximately 22 seconds before gently wicking away the excess solution using filter paper. To ensure thorough drying, the stained grids were stored in a grid box overnight. Afterward, TEM imaging was performed using a Tecnai TF-20 microscope (FEI Company, Hillsboro, OR) operating at 200kV.

### Scanning electron microscopy (SEM)

The BARA with SARS-CoV-2 virions captured on the surface with recombinant ACE2 was dehydrated with increasing ethanol concentrations (70, 85, 95, and 100 % (v/v)) for 5 min each. Lastly, the BARA was immersed in hexamethyldisilazane (HMDS, Sigma-Aldrich) for 10 min and air-dried overnight. The samples were imaged using an Apreo 2 SEM (FEI Company, Hillsboro, OR).

### Flow cytometry

SARS-CoV-2 virions were stained with antibodies targeting the spike glycoprotein and molecular beacons targeting the nucleocapsid-encoding RNA (**Table S10**), according to the previous strategies. MLV virions and PBS were utilized as a negative control and were stained following the same protocol. The samples were then imaged using ImageStream ® ^X^ mark II (MilliporeSigma, Burlington, MA, USA).

## Data Availability

All data produced in the present study are available upon reasonable request to the authors.

## Acknowledgments

We acknowledge all the patients and healthy volunteers participating in this study. We thank BEI Resources for providing the microorganism for cross-reactivity and microbial interference tests. We thank Dr. Eliah Aronoff-Spencer, Dr. Aaron Carlin, and Dr. Yves Theriault for preparing and providing the different SARS-CoV-2 variants used in this study. We thank the RADx-Radical Network for fruitful discussions during the development of this work. We thank Dr. Xiaokui Mo for helpful advice on the statistical tests. Electron microscopy was performed at the Center for Electron Microscopy and Analysis (CEMAS) at The Ohio State University. High-resolution flow cytometry was performed at Flow Cytometry Shared Resource (FCSR) at The Ohio State University, Comprehensive Cancer Center. We thank Anthony S. Baker and Courtney Fleming at the Health Sciences Library Medical Visuals at The Ohio State University for the illustration artwork.

## Author contributions

E.R. conceived the idea. K.T.N., E.R. developed the BARA technology. K.T.N., L.J.L., and E.R. designed the study. K.T.N., X.Y.R., L.T.H.N., X.W., and E.R. analyzed the data. K.T.N., X.Y.R., E.R., and S.M.M. prepared the figures and wrote the manuscript with input from all authors. K.T.N., L.T.H.N., and M.J.Y. performed the BARA experiments, qRT-PCR, TEM, flow cytometry, and biomarker characterization. K.J.K. designed the molecular beacons and prepared the biochips. X.W. and X.Y.R. developed the custom-built algorithm for image analysis. C.-L.C. performed on-chip qRT-PCR analysis. K.T.N., L.T.H.N., and M.J.Y. purified virions, EVs using qEV. A.F.P. provided purification protocols. M.J.Y. and X.Y.R. performed TRPS measurements. J.D.-R. obtained healthy donor blood. K.S., S.F., I.L., K.W., P.P., H.G.K., J.D.G., and J.R.H. provided clinical samples, Ct results, and patient clinical information. H.L. and X.W. assisted with biochip preparation. E.R. supervised the project and edited the manuscript. All authors have read and approved the final manuscript.

## Data and materials availability

All data needed to evaluate the conclusions in the paper are present in the paper and/or the Supplementary Materials. Additional data related to this paper may be requested from the authors.

## References

1. K. E. Jones et al., Global trends in emerging infectious diseases. Nature 451, 990–993 (2008).

2. R. K. Plowright et al., Pathways to zoonotic spillover. Nat Rev Microbiol 15, 502–510 (2017).

3. W. B. Karesh et al., Ecology of zoonoses: natural and unnatural histories. Lancet 380, 1936–1945 (2012).

4. I. Y. Chu, P. Alam, H. J. Larson, L. Lin, Social consequences of mass quarantine during epidemics: a systematic review with implications for the COVID-19 response. J Travel Med 27 (2020).

5. N. Delassalle, M. Cavaciuti, Psychological Distress and COVID-19: Evidence-Based Interventions for Frontline Health Care Workers-A Literature Review. Dimens Crit Care Nurs 42, 53–62 (2023).

6. J. J. V. Bavel et al., Using social and behavioural science to support COVID-19 pandemic response. Nat Hum Behav 4, 460–471 (2020).

7. A. L. Pedrosa et al., Emotional, Behavioral, and Psychological Impact of the COVID-19 Pandemic. Front Psychol 11, 566212 (2020).

8. World Health Organization. WHO Coronavirus (COVID-19) Dashboard. Available online: https://covid19.who.int/ (accessed on 7 March 2023).

9. M. S. Shiels et al., Racial and Ethnic Disparities in Excess Deaths During the COVID-19 Pandemic, March to December 2020. Ann Intern Med 174, 1693–1699 (2021).

10. E. C. Holmes, COVID-19-lessons for zoonotic disease. Science 375, 1114–1115 (2022).

11. C. J. Carlson et al., Climate change increases cross-species viral transmission risk. Nature 607, 555–562 (2022).

12. B. H. Bird, J. A. K. Mazet, Detection of Emerging Zoonotic Pathogens: An Integrated One Health Approach. Annu Rev Anim Biosci 6, 121–139 (2018).

13. Q. X. Long et al., Clinical and immunological assessment of asymptomatic SARS-CoV-2 infections. Nat Med 26, 1200–1204 (2020).

14. N. Sethuraman, S. S. Jeremiah, A. Ryo, Interpreting Diagnostic Tests for SARS-CoV-2. JAMA 323, 2249–2251 (2020).

15. D. Jacofsky, E. M. Jacofsky, M. Jacofsky, Understanding Antibody Testing for COVID-19. J Arthroplasty 35, S74–S81 (2020).

16. B. Lou et al., Serology characteristics of SARS-CoV-2 infection after exposure and post-symptom onset. Eur Respir J 56 (2020).

17. A. Tahamtan, A. Ardebili, Real-time RT-PCR in COVID-19 detection: issues affecting the results. Expert Rev Mol Diagn 20, 453–454 (2020).

18. M. Teymouri et al., Recent advances and challenges of RT-PCR tests for the diagnosis of COVID-19. Pathol Res Pract 221, 153443 (2021).

19. Y. Zhang, R. Garner, S. Salehi, M. Rocca, D. Duncan, Molecular and antigen tests, and sample types for diagnosis of COVID-19: a review. Future Virol 17, 675–685 (2022).

20. Y. Wu et al., Detection of extracellular RNAs in cancer and viral infection via tethered cationic lipoplex nanoparticles containing molecular beacons. Anal Chem 85, 11265–11274 (2013).

21. C. Hepp et al., Viral detection and identification in 20 min by rapid single-particle fluorescence in-situ hybridization of viral RNA. Sci Rep 11, 19579 (2021).

22. Y. Zhang, J. T. Bahns, Q. Jin, R. Divan, L. Chen, Toward the detection of single virus particle in serum. Anal Biochem 356, 161–170 (2006).

23. N. Shiaelis et al., Virus Detection and Identification in Minutes Using Single-Particle Imaging and Deep Learning. ACS Nano 17, 697–710 (2023).

24. K. Watanabe, H. Y. Wu, J. Xavier, L. T. Joshi, F. Vollmer, Single Virus Detection on Silicon Photonic Crystal Random Cavities. Small 18, e2107597 (2022).

25. E. Nolte-’t Hoen, T. Cremer, R. C. Gallo, L. B. Margolis, Extracellular vesicles and viruses: Are they close relatives? Proc Natl Acad Sci U S A 113, 9155–9161 (2016).

26. N. Dhiman, R. Awasthi, B. Sharma, H. Kharkwal, G. T. Kulkarni, Lipid Nanoparticles as Carriers for Bioactive Delivery. Front Chem 9, 580118 (2021).

27. E. Willms, C. Cabañas, I. Mäger, M. J. A. Wood, P. Vader, Extracellular Vesicle Heterogeneity: Subpopulations, Isolation Techniques, and Diverse Functions in Cancer Progression. Front Immunol 9, 738 (2018).

28. C. Y. Chiang, C. Chen, Toward characterizing extracellular vesicles at a single-particle level. J Biomed Sci 26, 9 (2019).

29. S. Dechantsreiter et al., Heterogeneity in extracellular vesicle secretion by single human macrophages revealed by super-resolution microscopy. J Extracell Vesicles 11, e12215 (2022).

30. M. S. Panagopoulou, A. W. Wark, D. J. S. Birch, C. D. Gregory, Phenotypic analysis of extracellular vesicles: a review on the applications of fluorescence. J Extracell Vesicles 9, 1710020 (2020).

31. X. Y. Rima et al., Microfluidic harvesting of breast cancer tumor spheroid-derived extracellular vesicles from immobilized microgels for single-vesicle analysis. Lab Chip 22, 2502–2518 (2022).

32. J. Zhou et al., High-throughput single-EV liquid biopsy: Rapid, simultaneous, and multiplexed detection of nucleic acids, proteins, and their combinations. Sci Adv 6 (2020).

33. C. Han et al., Single-vesicle imaging and co-localization analysis for tetraspanin profiling of individual extracellular vesicles. J Extracell Vesicles 10, e12047 (2021).

34. K. Lee et al., Multiplexed Profiling of Single Extracellular Vesicles. ACS Nano 12, 494–503 (2018).

35. J. D. Spitzberg et al., Multiplexed analysis of EV reveals specific biomarker composition with diagnostic impact. Nat Commun 14, 1239 (2023).

36. K. Breitwieser et al., Detailed Characterization of Small Extracellular Vesicles from Different Cell Types Based on Tetraspanin Composition by ExoView R100 Platform. Int J Mol Sci 23 (2022).

37. L. T. H. Nguyen et al., An immunogold single extracellular vesicular RNA and protein (J Extracell Vesicles 11, e12258 (2022).

38. D. L. Floyd, J. R. Ragains, J. J. Skehel, S. C. Harrison, A. M. van Oijen, Single-particle kinetics of influenza virus membrane fusion. Proc Natl Acad Sci U S A 105, 15382–15387 (2008).

39. S. Ivanchenko et al., Dynamics of HIV-1 assembly and release. PLoS Pathog 5, e1000652 (2009).

40. V. Baumgärtel et al., Live-cell visualization of dynamics of HIV budding site interactions with an ESCRT component. Nat Cell Biol 13, 469–474 (2011).

41. A. C. Walls et al., Structure, Function, and Antigenicity of the SARS-CoV-2 Spike Glycoprotein. Cell 181, 281–292.e286 (2020).

42. M. Letko, A. Marzi, V. Munster, Functional assessment of cell entry and receptor usage for SARS-CoV-2 and other lineage B betacoronaviruses. Nat Microbiol 5, 562–569 (2020).

43. J. Zhang et al., Engineering a Single Extracellular Vesicle Protein and RNA Assay (^siEV^PRA) via In Situ Fluorescence Microscopy in a UV Micropatterned Array. bioRxiv, 2022.2008.2005.502995 (2022).

44. W. T. Harvey et al., SARS-CoV-2 variants, spike mutations and immune escape. Nat Rev Microbiol 19, 409–424 (2021).

45. K. Ota et al., Detection of SARS-CoV-2 using qRT-PCR in saliva obtained from asymptomatic or mild COVID-19 patients, comparative analysis with matched nasopharyngeal samples. PLoS One 16, e0252964 (2021).

46. J. Hellewell et al., Estimating the effectiveness of routine asymptomatic PCR testing at different frequencies for the detection of SARS-CoV-2 infections. BMC Med 19, 106 (2021).

47. T. Thaweethai et al., Development of a Definition of Postacute Sequelae of SARS-CoV-2 Infection. JAMA 329, 1934–1946 (2023).

48. Y. Mina et al., Deep Phenotyping of Neurologic Postacute Sequelae of SARS-CoV-2 Infection. Neurol Neuroimmunol Neuroinflamm 10 (2023).

49. Y. Xie, E. Xu, B. Bowe, Z. Al-Aly, Long-term cardiovascular outcomes of COVID-19. Nat Med 28, 583–590 (2022).

50. H. E. Davis, L. McCorkell, J. M. Vogel, E. J. Topol, Long COVID: major findings, mechanisms and recommendations. Nat Rev Microbiol 21, 133–146 (2023).

51. J. Kowal et al., Proteomic comparison defines novel markers to characterize heterogeneous populations of extracellular vesicle subtypes. Proc Natl Acad Sci U S A 113, E968–977 (2016).

52. C. S. Greenberg, C. C. Miraglia, F. R. Rickles, M. A. Shuman, Cleavage of blood coagulation factor XIII and fibrinogen by thrombin during in vitro clotting. J Clin Invest 75, 1463–1470 (1985).

53. A. D. Proal, M. B. VanElzakker, Long COVID or Post-acute Sequelae of COVID-19 (PASC): An Overview of Biological Factors That May Contribute to Persistent Symptoms. Front Microbiol 12, 698169 (2021).

54. L. A. Nikolai, C. G. Meyer, P. G. Kremsner, T. P. Velavan, Asymptomatic SARS Coronavirus 2 infection: Invisible yet invincible. Int J Infect Dis 100, 112–116 (2020).

55. K. A. Walsh et al., SARS-CoV-2 detection, viral load and infectivity over the course of an infection. J Infect 81, 357–371 (2020).

56. K. F. Jarvis, J. B. Kelley, Temporal dynamics of viral load and false negative rate influence the levels of testing necessary to combat COVID-19 spread. Sci Rep 11, 9221 (2021).

57. I. Arevalo-Rodriguez et al., False-negative results of initial RT-PCR assays for COVID-19: A systematic review. PLoS One 15, e0242958 (2020).

58. D. S. Mouliou, K. I. Gourgoulianis, False-positive and false-negative COVID-19 cases: respiratory prevention and management strategies, vaccination, and further perspectives. Expert Rev Respir Med 15, 993–1002 (2021).

59. L. M. Kucirka, S. A. Lauer, O. Laeyendecker, D. Boon, J. Lessler, Variation in False-Negative Rate of Reverse Transcriptase Polymerase Chain Reaction-Based SARS-CoV-2 Tests by Time Since Exposure. Ann Intern Med 173, 262–267 (2020).

60. W. J. Wiersinga, A. Rhodes, A. C. Cheng, S. J. Peacock, H. C. Prescott, Pathophysiology, Transmission, Diagnosis, and Treatment of Coronavirus Disease 2019 (COVID-19): A Review. JAMA 324, 782–793 (2020).

61. H. Im et al., Label-free detection and molecular profiling of exosomes with a nano-plasmonic sensor. Nat Biotechnol 32, 490–495 (2014).

62. W. T. He et al., Virome characterization of game animals in China reveals a spectrum of emerging pathogens. Cell 185, 1117–1129.e1118 (2022).

63. D. Groff et al., Short-term and Long-term Rates of Postacute Sequelae of SARS-CoV-2 Infection: A Systematic Review. JAMA Netw Open 4, e2128568 (2021).

64. J. A. Frontera et al., Post-acute sequelae of COVID-19 symptom phenotypes and therapeutic strategies: A prospective, observational study. PLoS One 17, e0275274 (2022).

65. Y. Su et al., Multiple early factors anticipate post-acute COVID-19 sequelae. Cell 185, 881–895.e820 (2022).

66. Y. Su et al., Multi-Omics Resolves a Sharp Disease-State Shift between Mild and Moderate COVID-19. Cell 183, 1479–1495.e1420 (2020).

