## Supporting Information for "Integrated antigenic and nucleic acid detection in single virions and virion-infected host-derived extracellular vesicles"

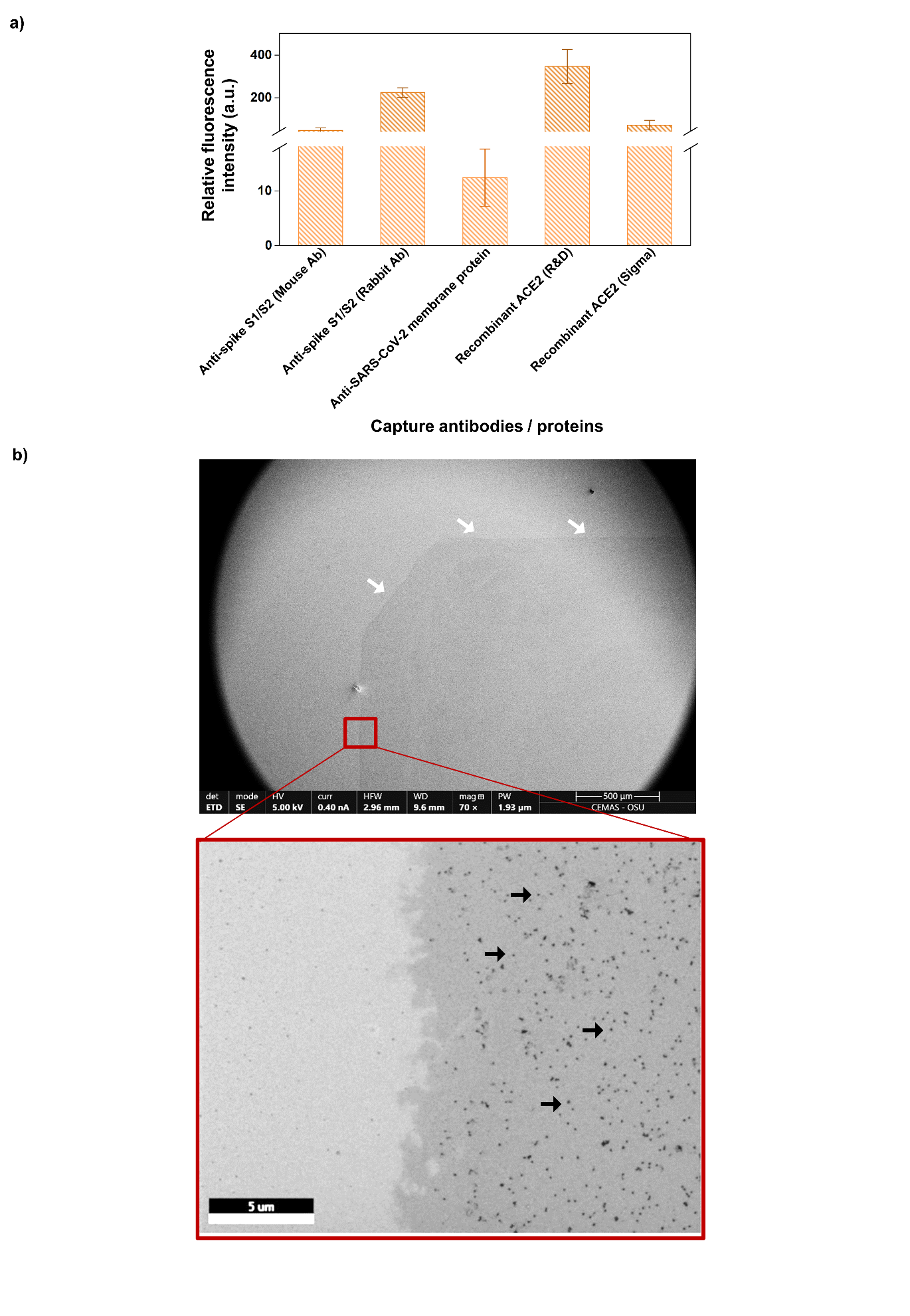

**Figure S1: Characterization and optimization of the BARA for detecting single SARS-CoV-2 virions. (a)** Single SARS-CoV-2 virions captured using different capture antibodies, including mouse-derived antibodies targeting the S1/S2 subunits of the spike glycoprotein, rabbit-derived antibodies targeting the S1/S2 subunits, antibodies targeting the SARS-CoV-2 membrane protein antibody, and recombinant ACE2 from R&D Systems and Sigma-Aldrich and detected for the spike glycoprotein (**Table S1;** n = 2, error bars indicate the standard deviation). **(b)** Scanning electron microscopy (SEM) images displaying single virions captured on the BARA, obtained through secondary electron detection. White arrows indicate the boundary between the bare gold surface and the antibody-coated inside the multiwell chamber area, where the antibody-coated area appears as a darker shade compared to the highly conductive gold surface. Black arrows indicate single SARS-CoV-2 virions captured on the antibody-coated surface.

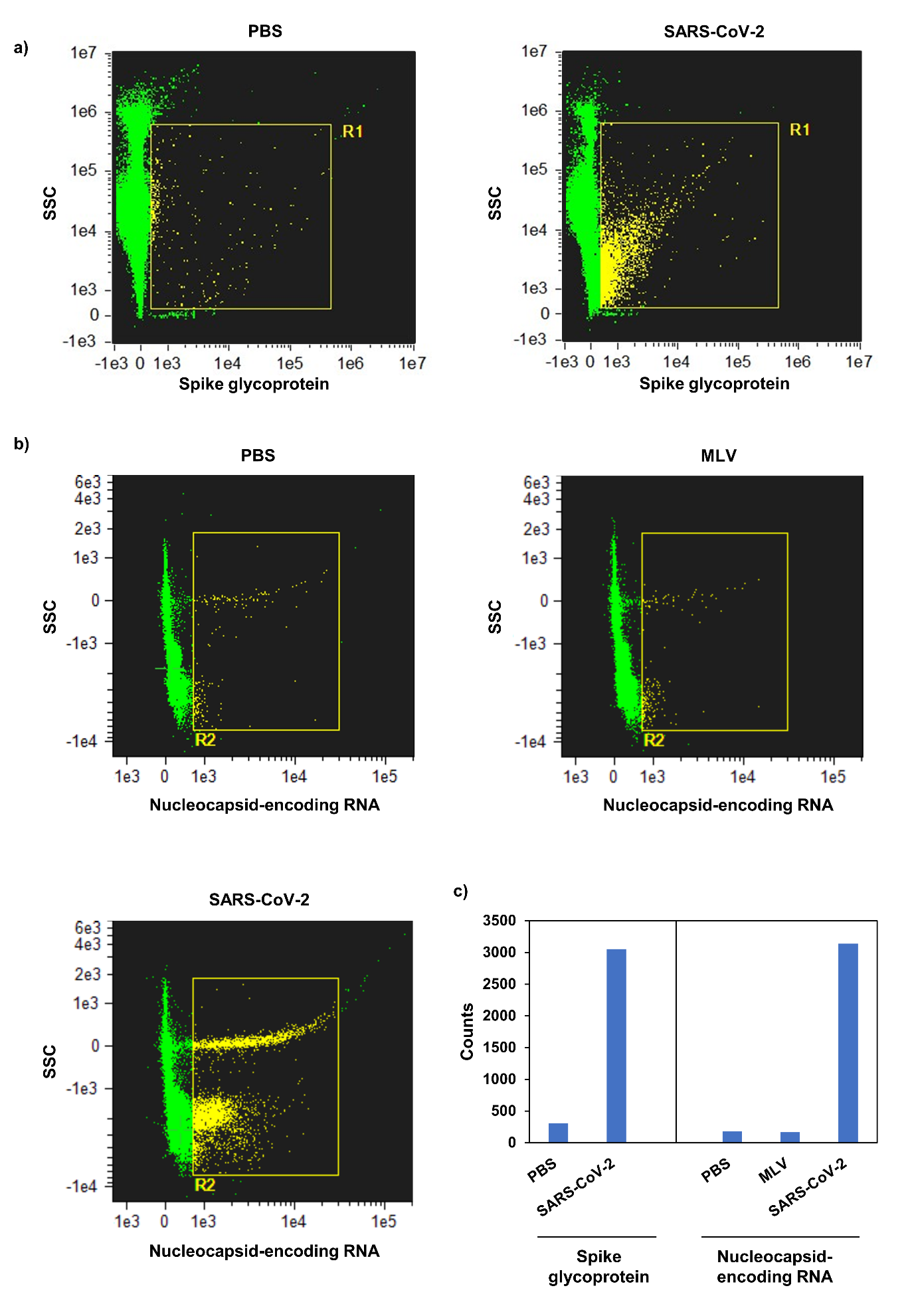

**Figure S2: Cross-validation of spike glycoprotein and nucleocapsid-encoding RNA detection in SARS-CoV-2 virions using high-resolution flow cytometry.** **(a)** Scatter plots depicting spike glycoprotein detection on virions via high-resolution flow cytometry. The R1 region highlights the subpopulation of SARS-CoV-2 virions with a high fluorescence signal for the spike glycoprotein. **(b)** Scatter plots showing the detection of SARS-CoV-2 nucleocapsid-encoding RNA using molecular beacons. The R2 region highlights the specific detection of the nucleocapsid-encoding RNA in SARS-CoV-2 virions compared to the murine leukemia virus (MLV) and negative control (phosphate-buffered saline; PBS). **(c)** Comparison of particle counts in the R1 and R2 regions for the respective samples. All samples were measured at E7 particles/mL.

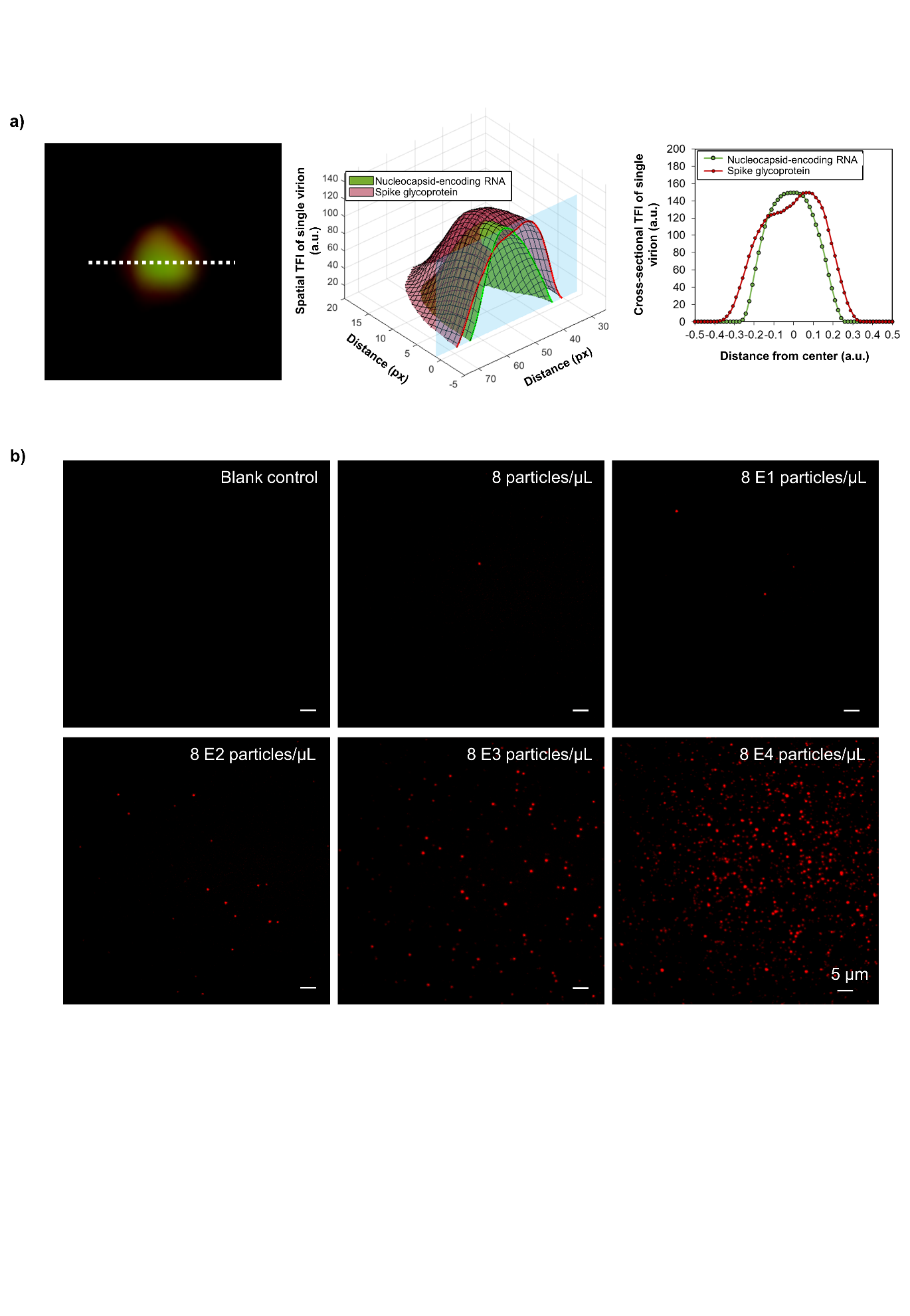

**Figure S3: Spatial characterization and dilution of a single SARS-CoV-2 virions with the BARA. (a)** Spatial expression of the spike glycoprotein and nucleocapsid-encoding RNA on a single SARS-CoV-2 virion, along with a cross-section of fluorescence intensity as a function of the *x – y* plane. **(b)** Representative total internal reflection fluorescence microscopy (TIRFM) images showing different concentrations of virion particles captured with the BARA.

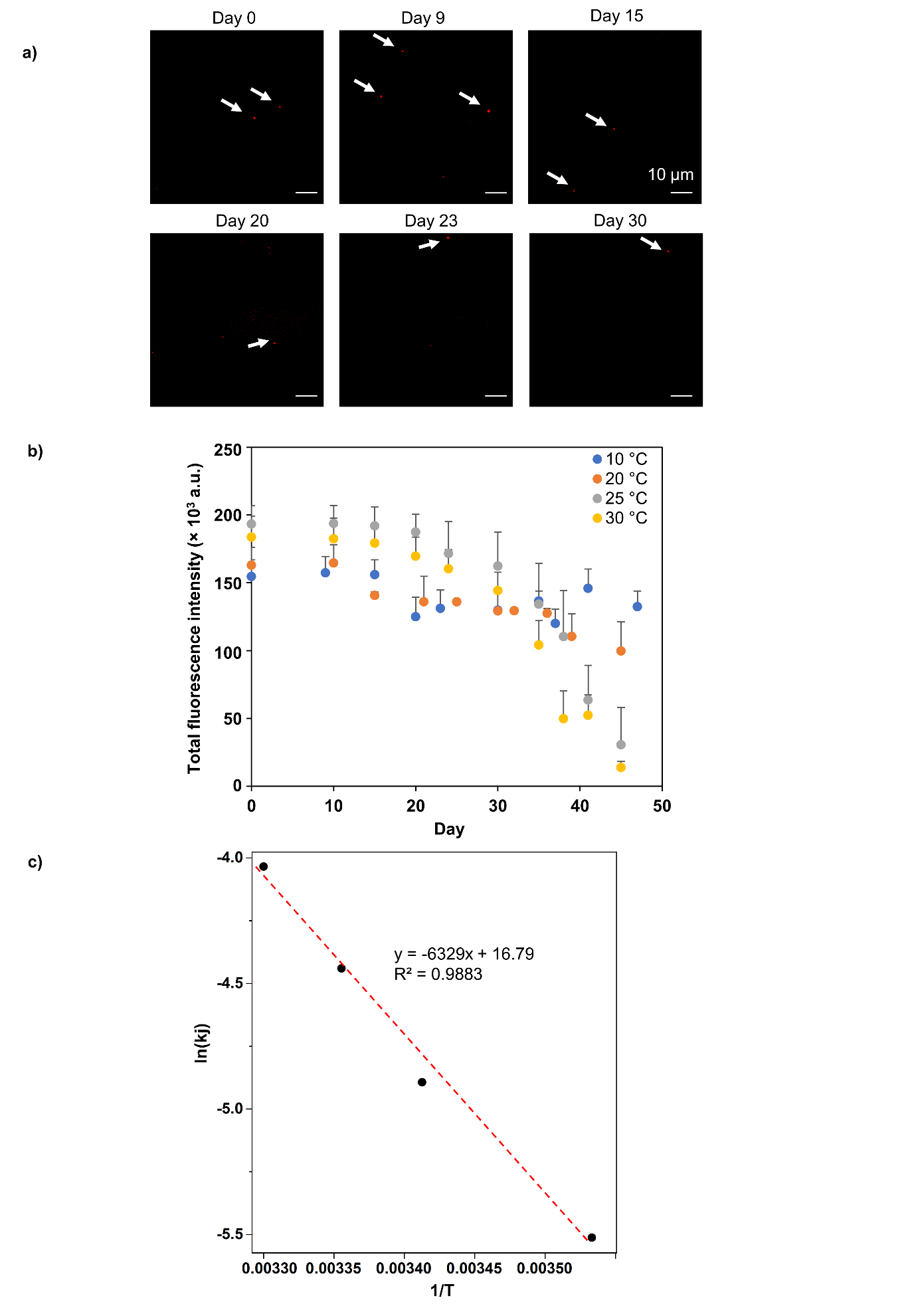

**Figure S4:** **Accelerated stability test for the BARA. (a)** TIRFM images acquired over multiple days at 10 °C. The white arrows indicate the individual SARS-CoV-2 virions detected. **(b)** Temperature-induced degradation of the biochip and its corresponding reagents measured over 47 days at four elevated temperatures (10, 20, 25, and 30 °C), with results obtained at ten different time points (N = 3, n = 5, error bars indicate the standard deviation) **(c)** Determination of the degradation rate constant of the biochip and its corresponding reagents at various temperatures using the Arrhenius equation and the methodology outlined in the Clinical and Laboratory Standards Institute (CLSI) EP25-A document. The calculated stability time (*t_stab_*) was 94.26 days at 4 °C for a 10% degradation.

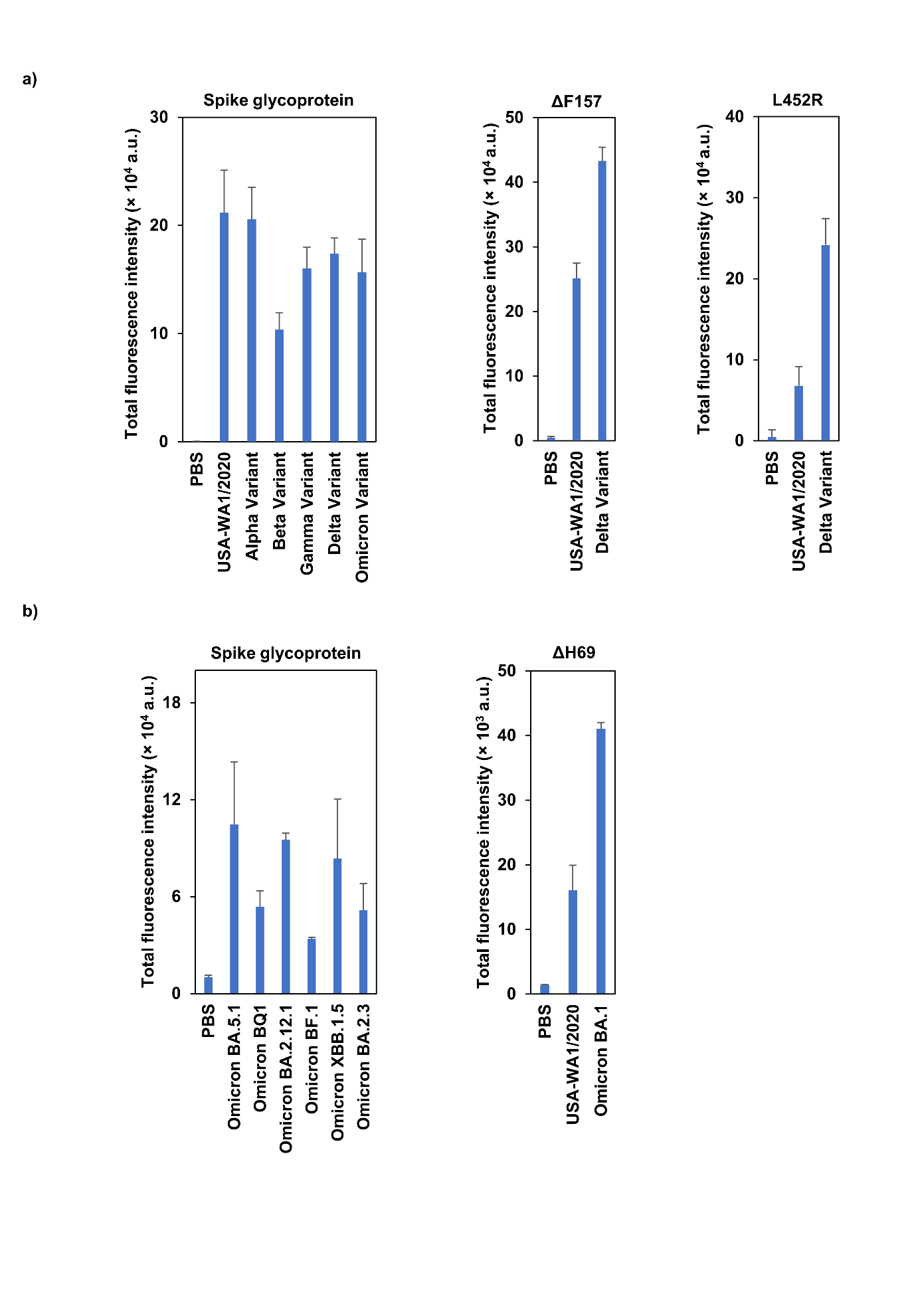

**Figure S5:** **Genetic mutations on single SARS-CoV-2 virions with the BARA.** **(a)** Total fluorescence intensities of spike glycoprotein detection across variants and expression of ΔF157 and L452R mutations specific to the delta variant (n = 2, error bars indicate the standard deviation). **(b)** Total fluorescence intensities of six Pango lineages of the omicron variant for the spike glycoprotein and molecular beacons targeting the ΔH69 mutation specific to the omicron variant (n = 2, error bars indicate the standard deviation).

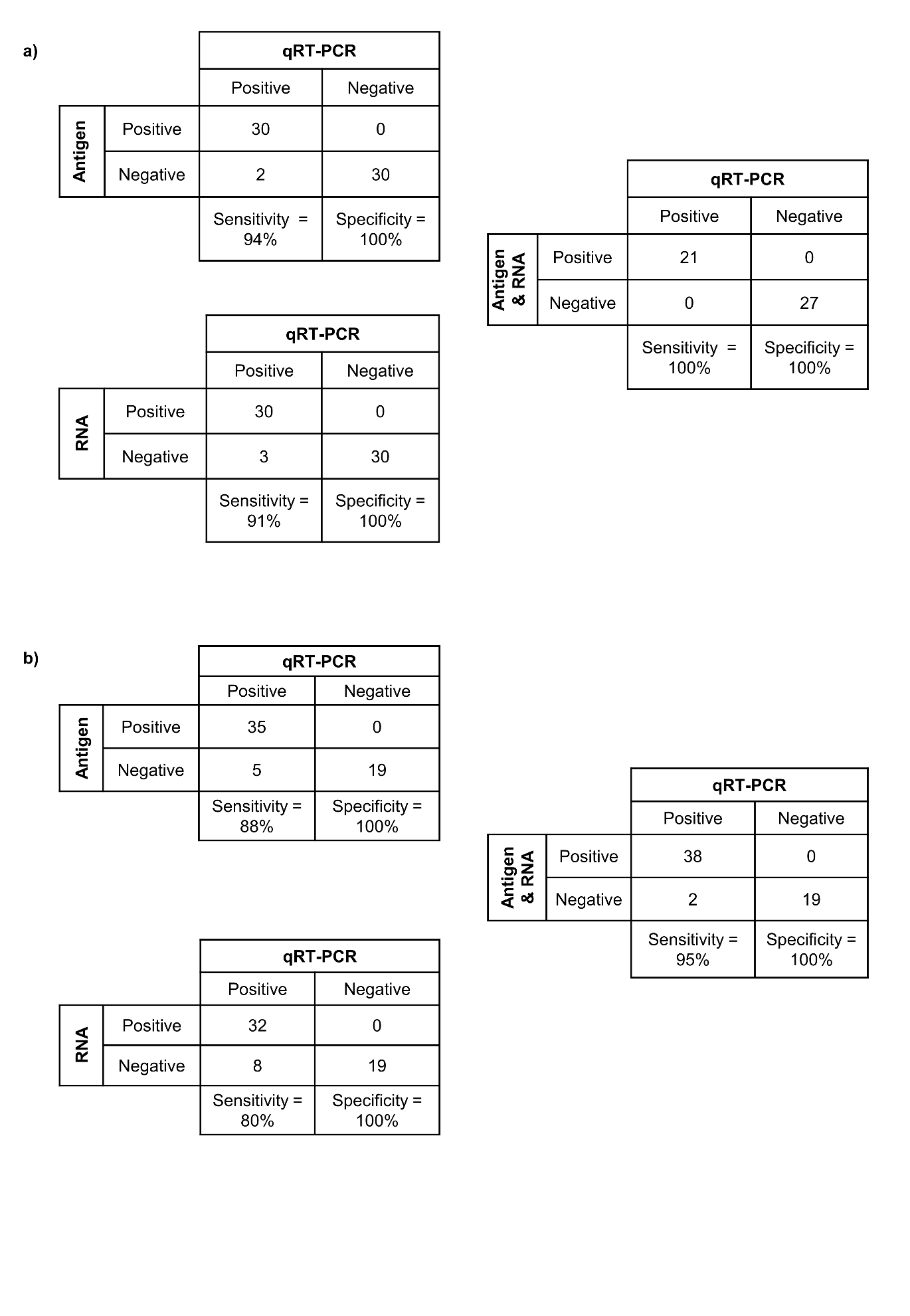

**Figure S6: Sensitivity and specificity for COVID-19 diagnoses in saliva and nasopharyngeal swab (NS) specimens.** **(a)** Sensitivity and specificity for spike glycoprotein, nucleocapsid-encoding RNA, and dual detection using saliva specimens. **(b)** Sensitivity and specificity for spike glycoprotein, nucleocapsid-encoding RNA, and dual detection using NS specimens.

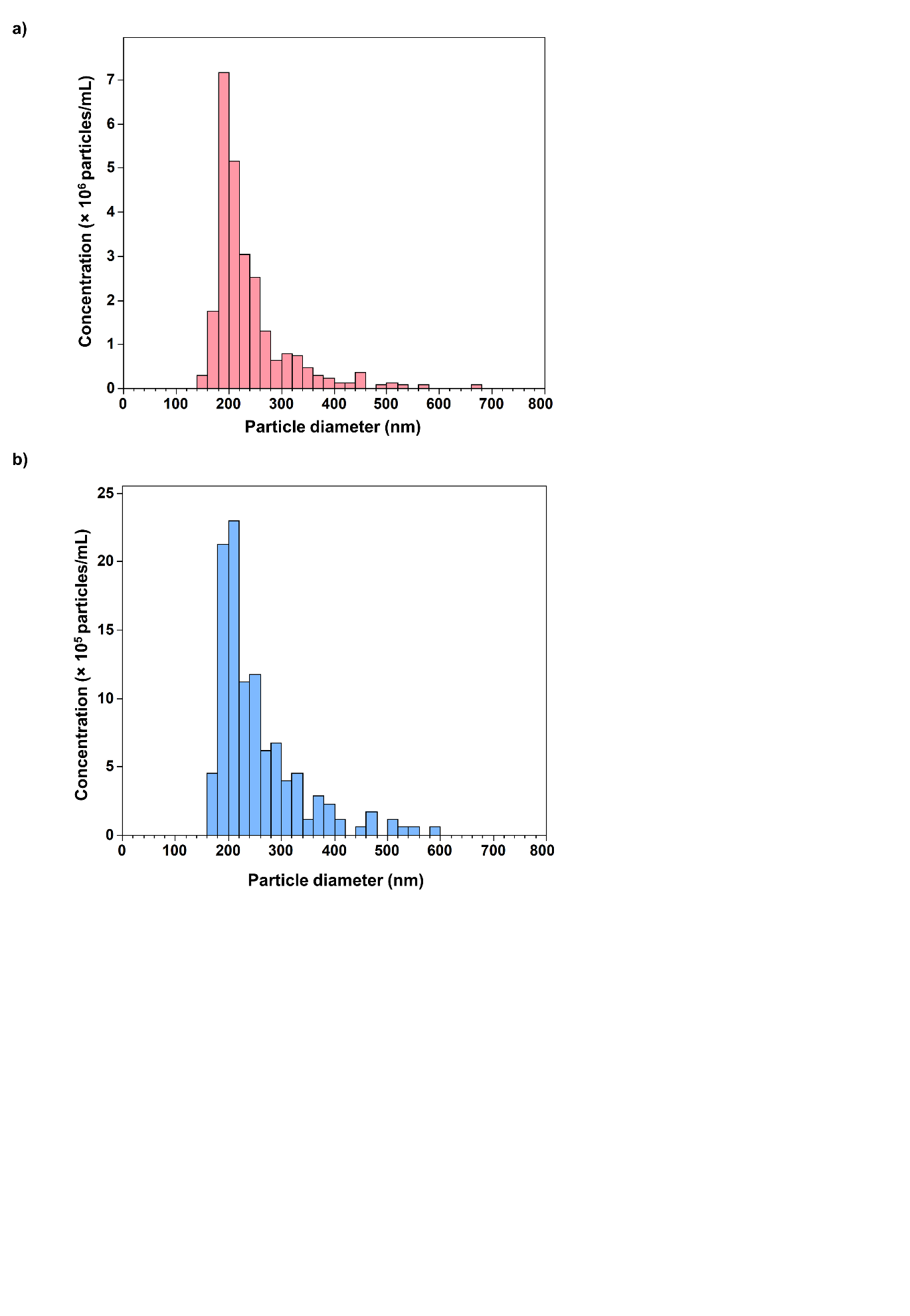

**Figure S7: Size distributions of SARS-CoV-2 virions and extracellular vesicles (EVs). (a)** Histogram representing the size distribution of extracellular vesicles (EVs) in healthy donor saliva specimens and purified by size-exclusion chromatography (SEC) measured using tunable resistive pulse sensing (TRPS). **(b)** Histogram displaying the size distribution of SARS-CoV-2 virions measured by TRPS.

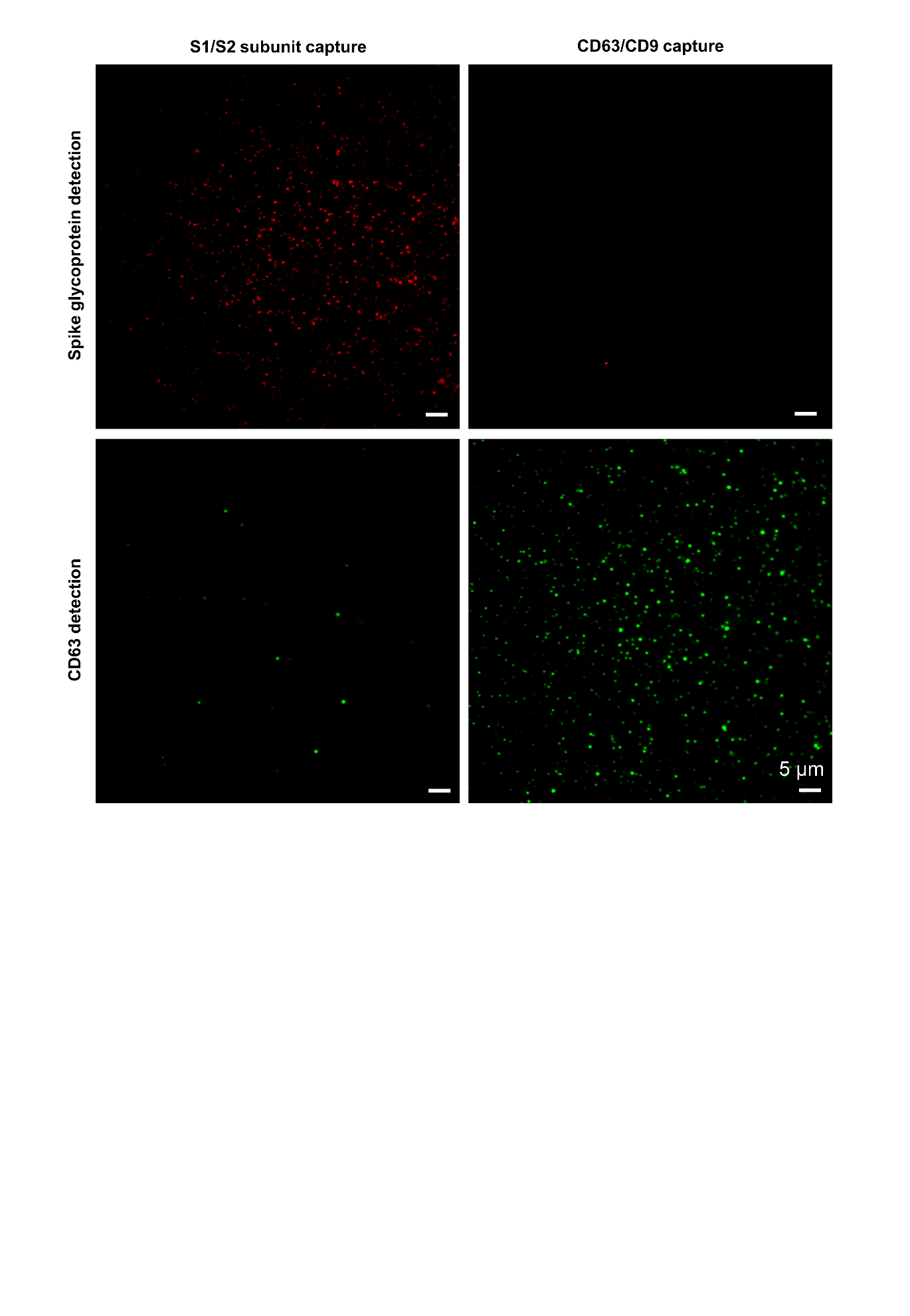

**Figure S8: SARS-CoV-2 virion and EVs sorting with the BARA.** TIRFM images illustrating specific capture and detection of subpopulations for SARS-CoV-2 virions and EVs from patient saliva samples. Antibodies targeting CD63/CD9 specifically captured CD63-expressing single EVs, whereas antibodies targeting the S1/S2 subunits of the spike glycoprotein enriched single SARS-CoV-2 virions expressing the spike glycoprotein.

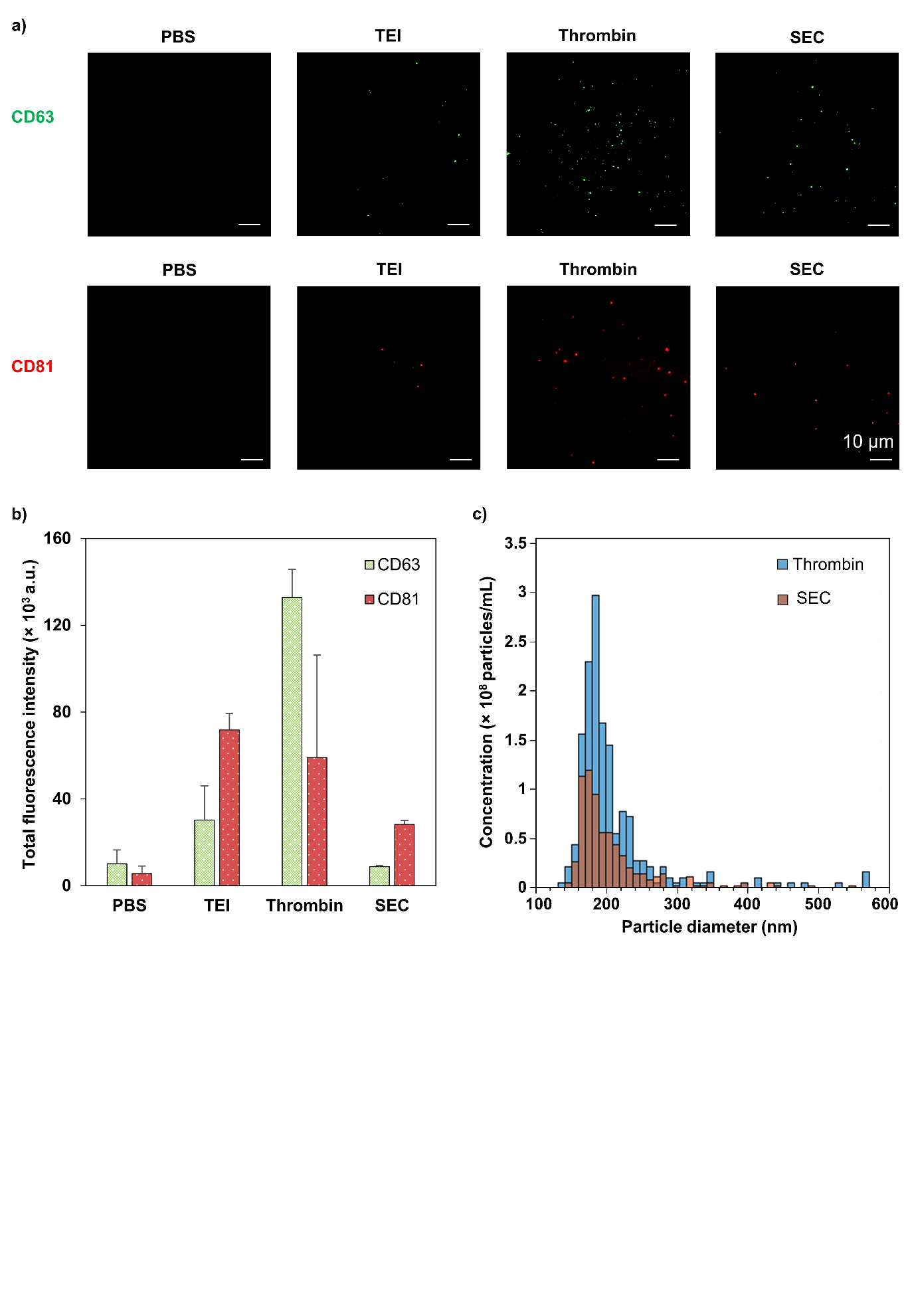

**Figure S9: EVs isolation from plasma. (a)** Representative TIRFM images showing EVs isolated from plasma using different isolation methods, including the total exosome isolation kit (TEI), thrombin, and SEC. **(b)** Evaluation of CD63^+^ and CD81^+^ EVs isolated from plasma of a healthy donor using the various isolation methods (n = 2, error bars indicate the standard deviation). **(c)** Comparison of TRPS analysis between plasma samples treated with thrombin and SEC purification.

**Table S1: List of antibodies and proteins used for virion and EV capture and detection.**

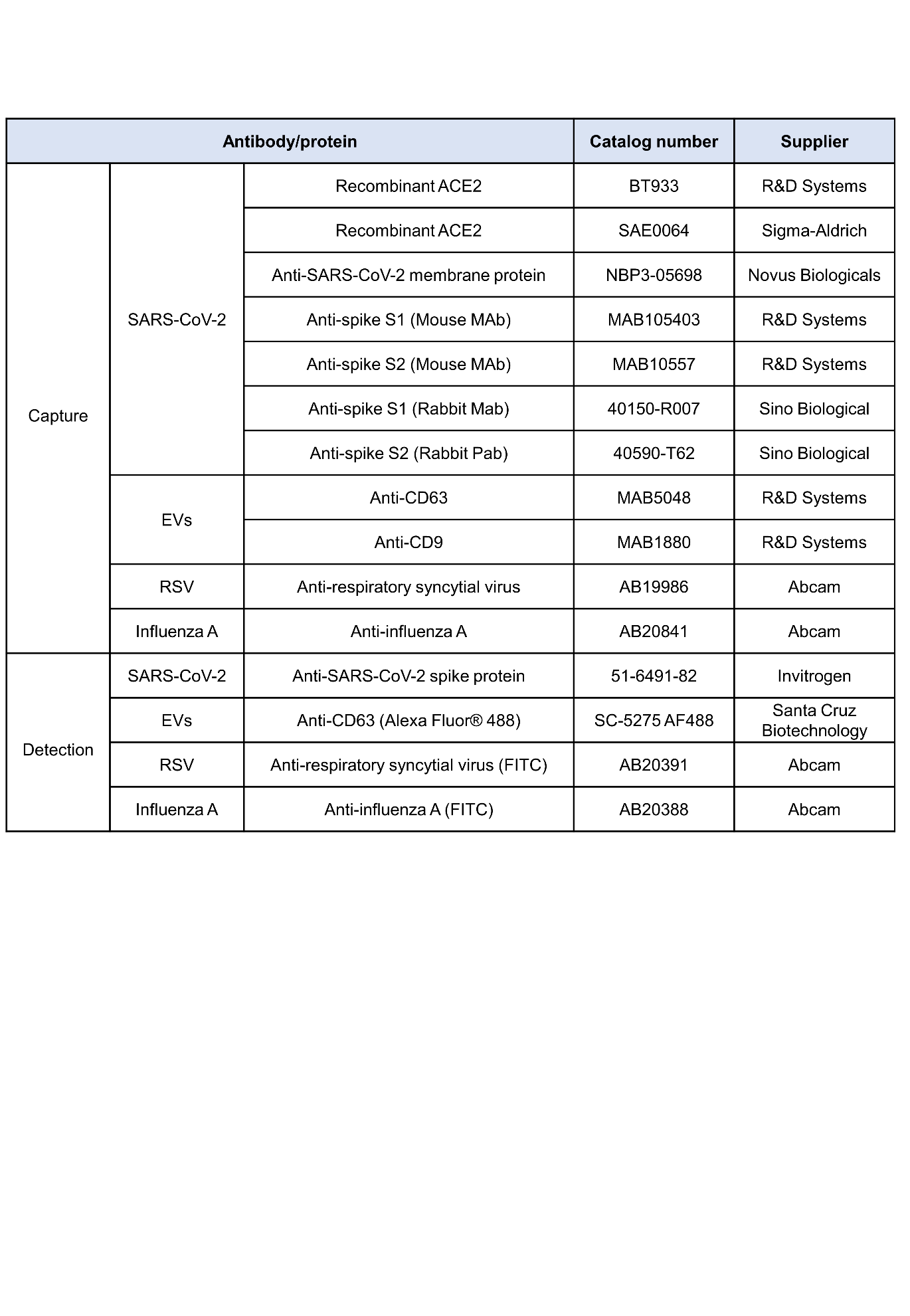

**Table S2: Testing concentrations for potential cross-reactive microorganisms.**

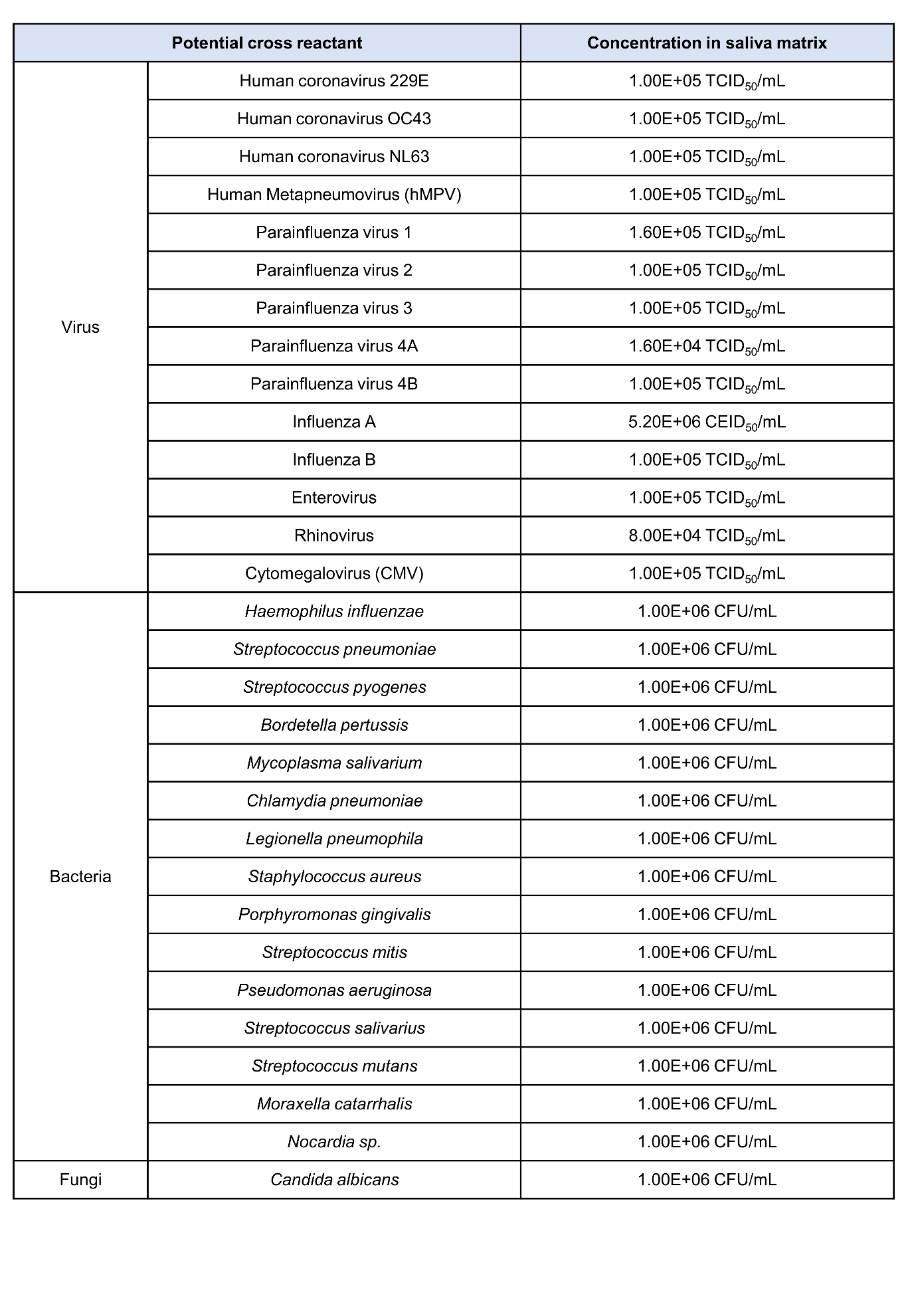

**Table S3: Testing concentrations for potential endogenous and exogenous substances.**

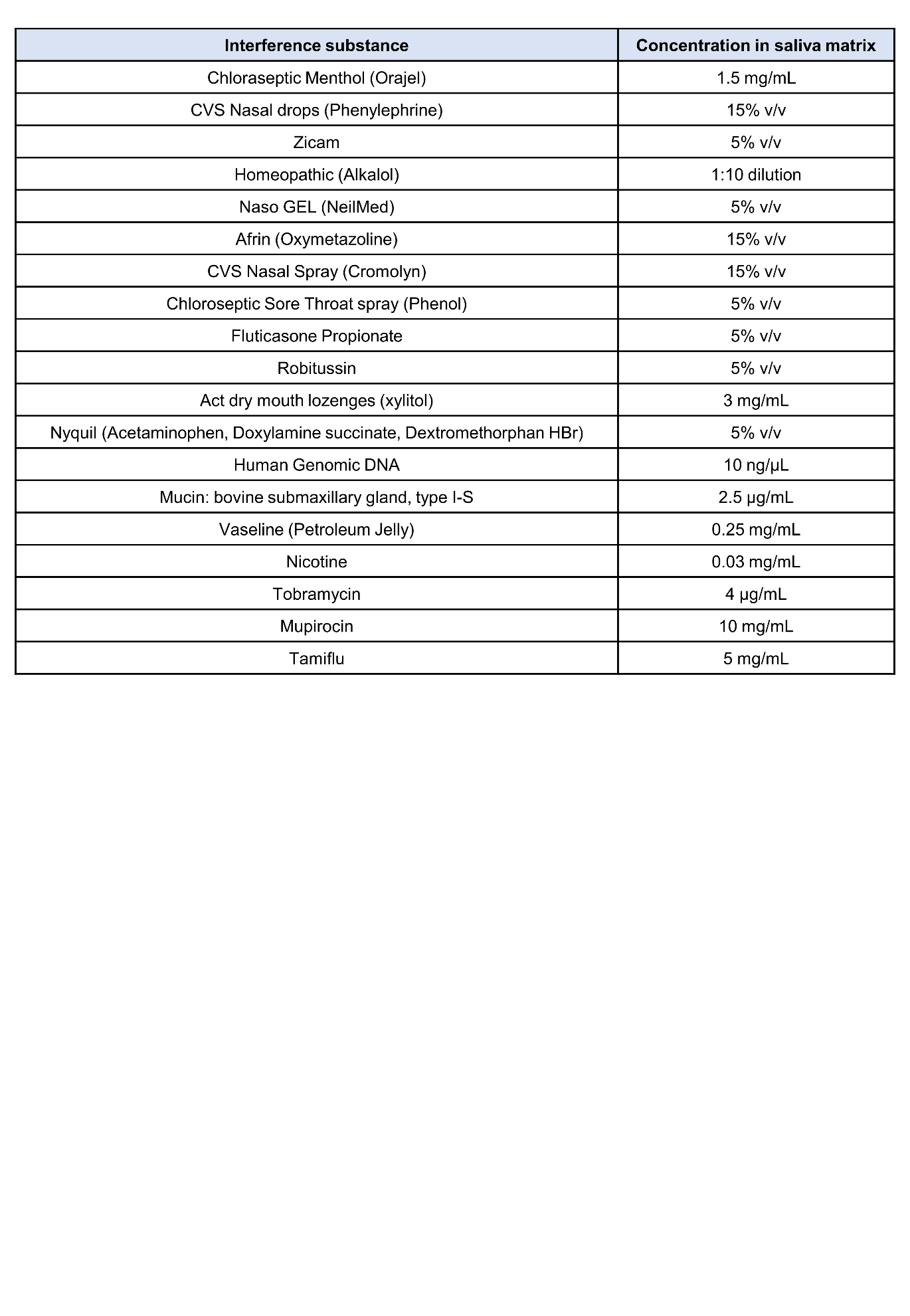

**Table S4:** **Accelerated stability test for the BARA at three times the limit of detection (LoD).**

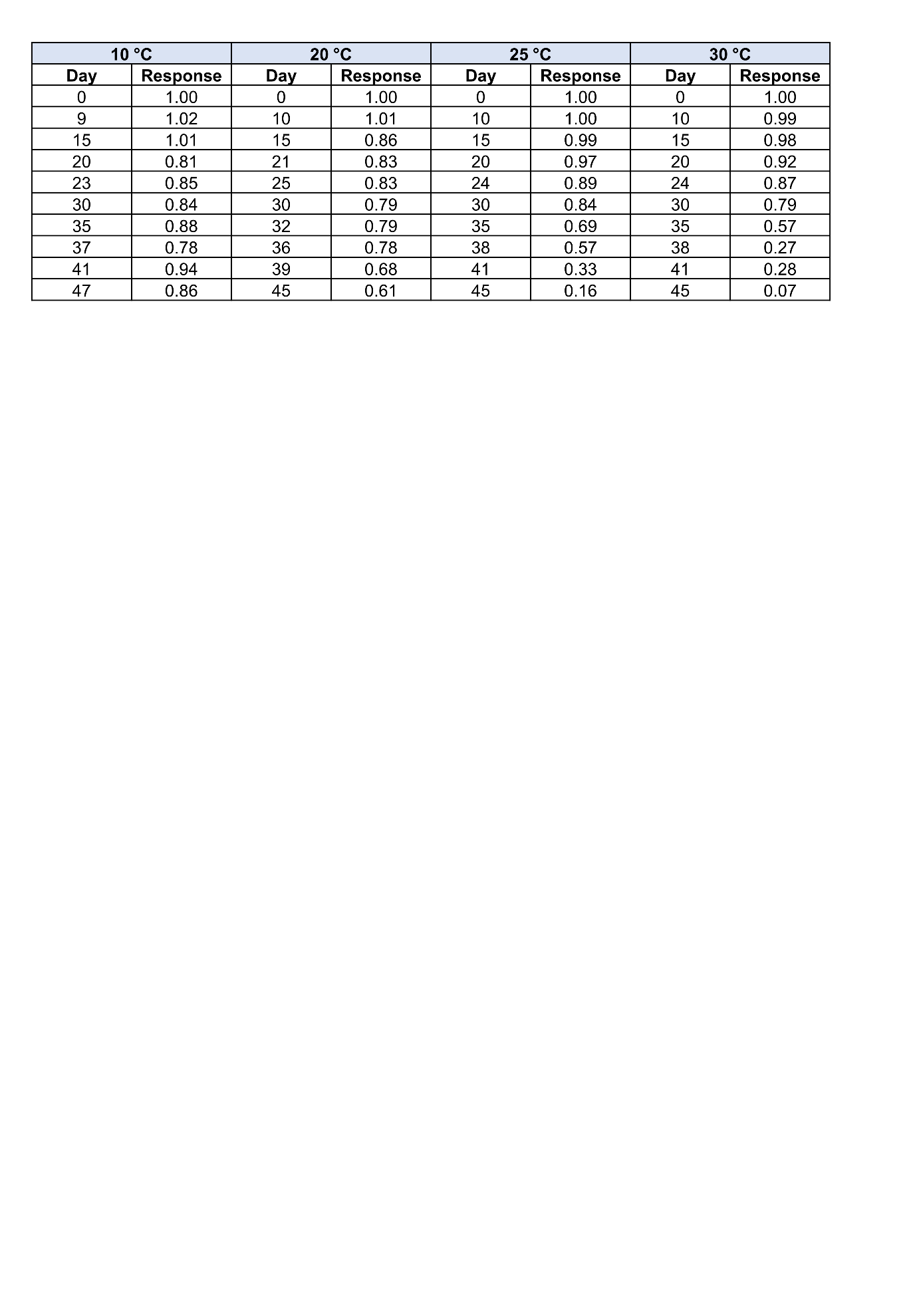

**Table S5: Degradation rate constants (*kj*) at different temperatures (*Tj*).**

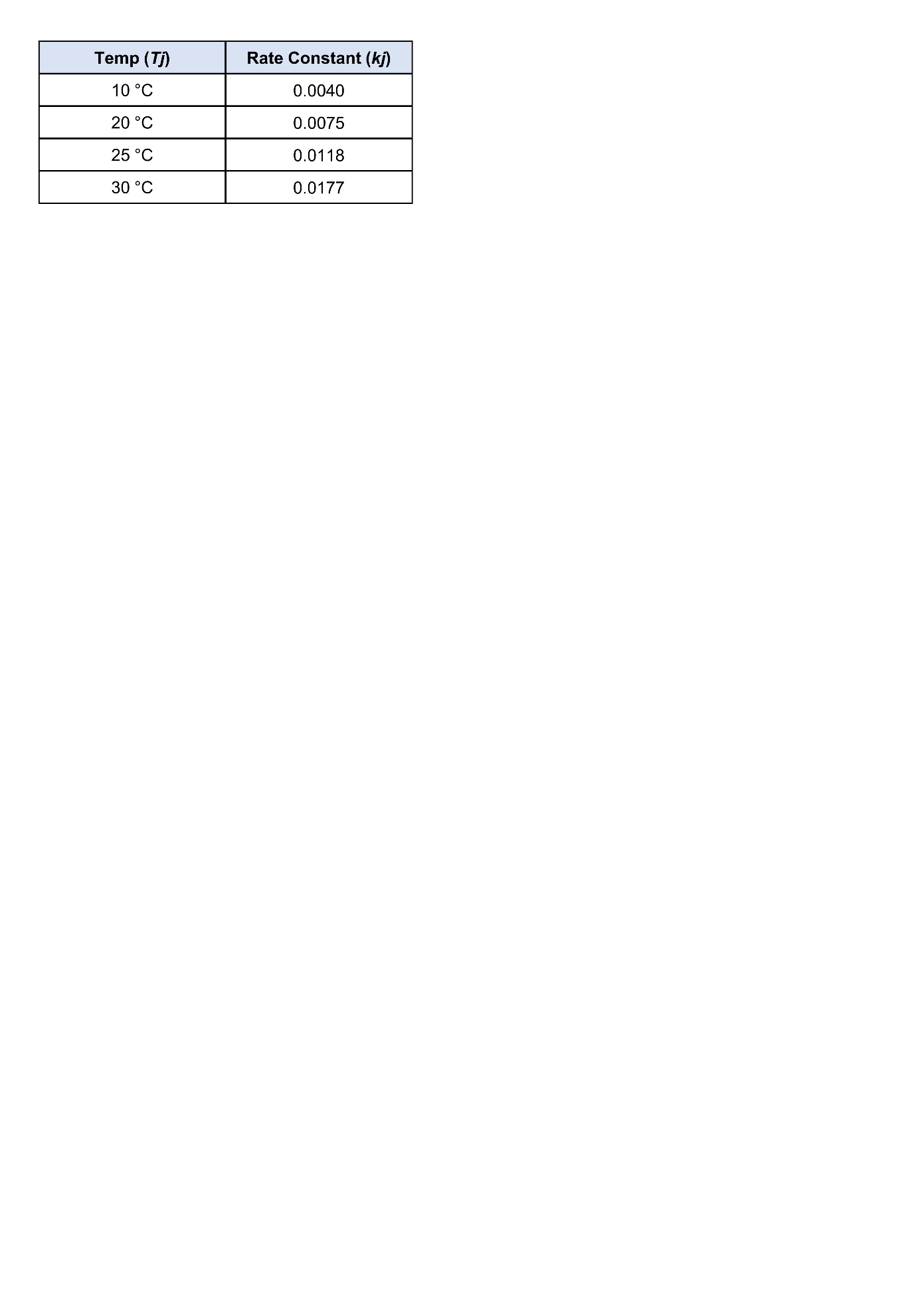

**Table S6: Detailed information on the patients and healthy donors enrolled for saliva collection.**

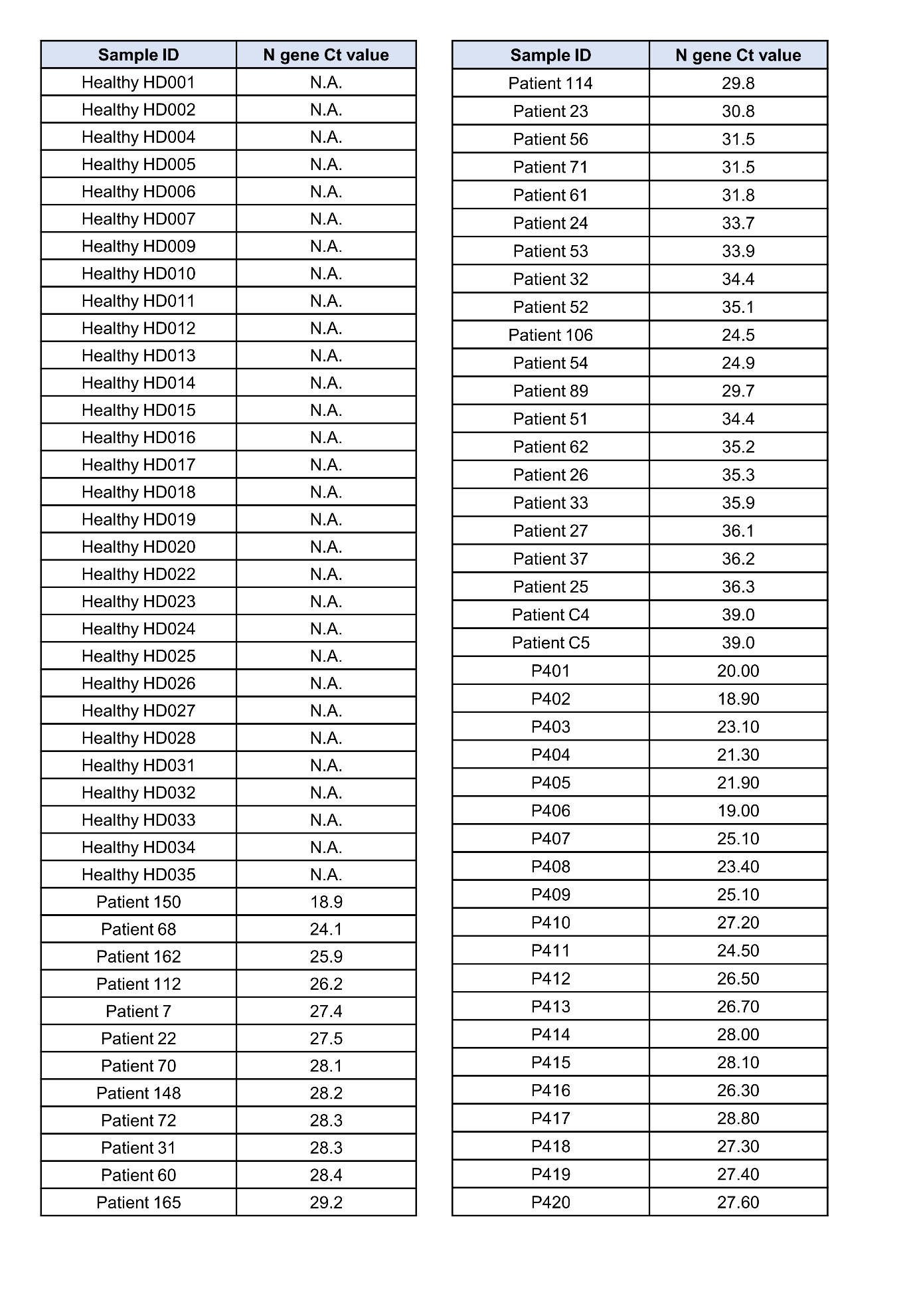

**Table S7: Detailed information on the patients and healthy donors enrolled for NS collection.**

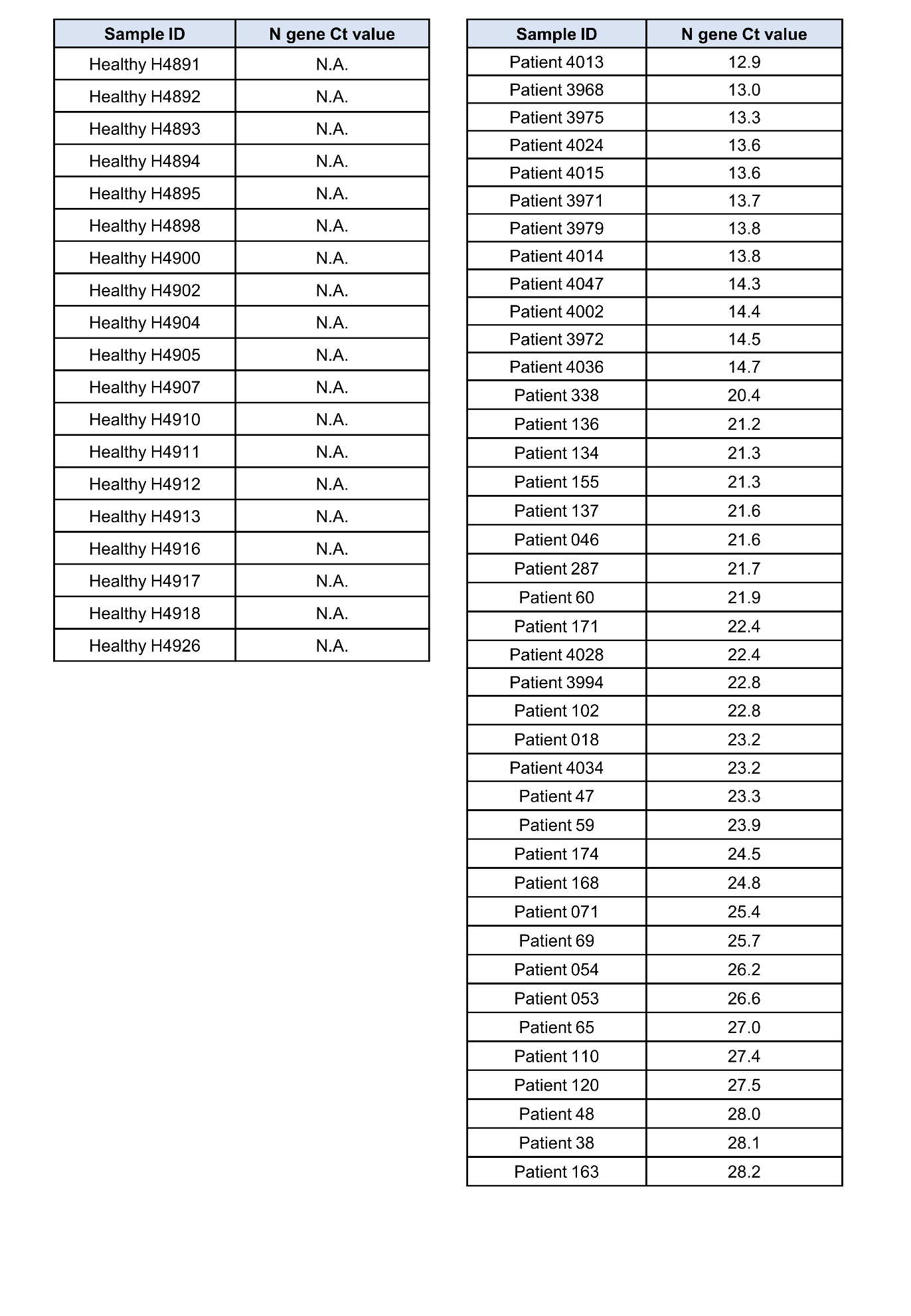

**Table S8: Detailed information on the patients enrolled for the post-acute sequelae SARS-CoV-2 infection (PASC) study.**

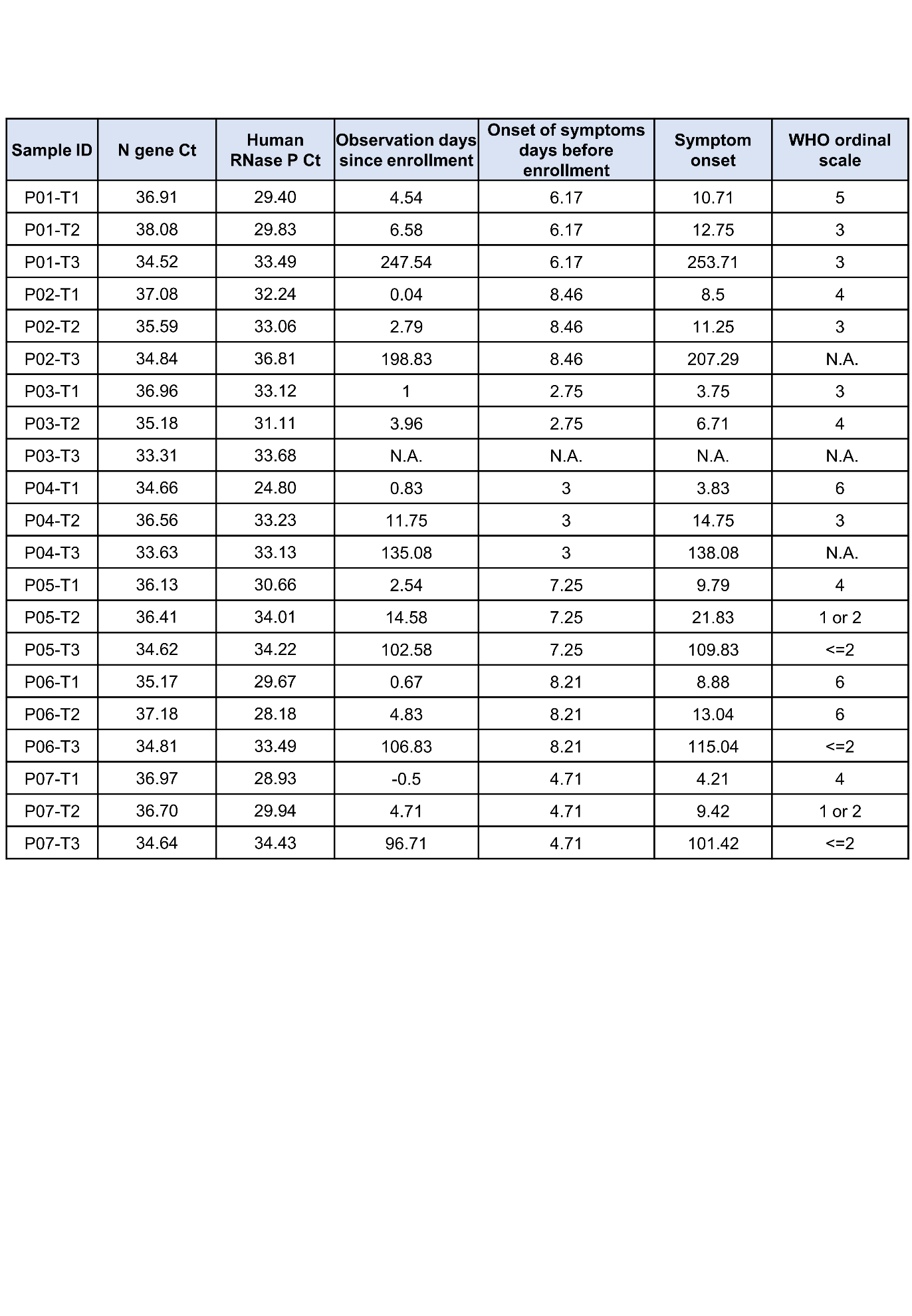

**Table S9:** **Symptom information for the PASC patients enrolled in the study.**

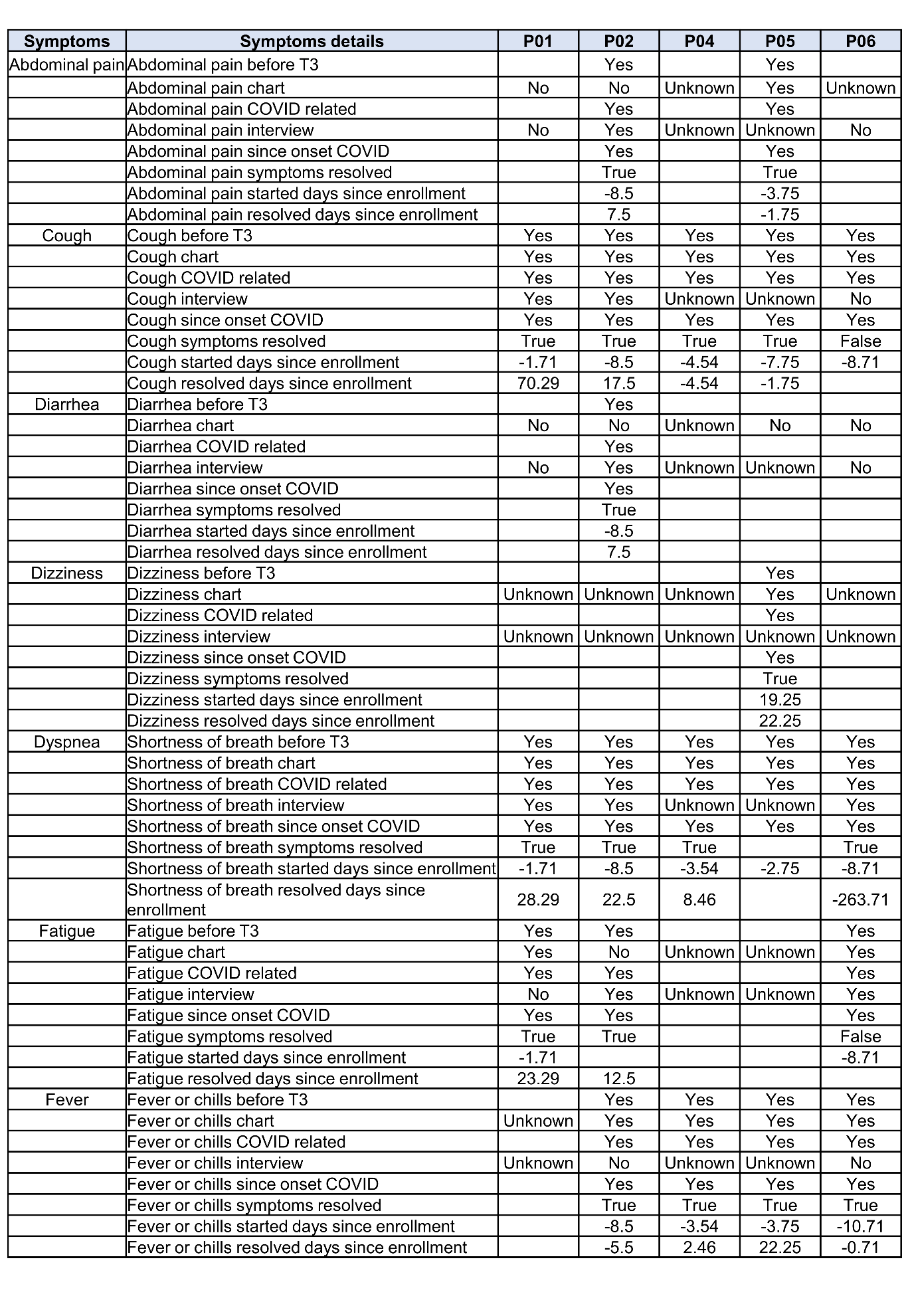

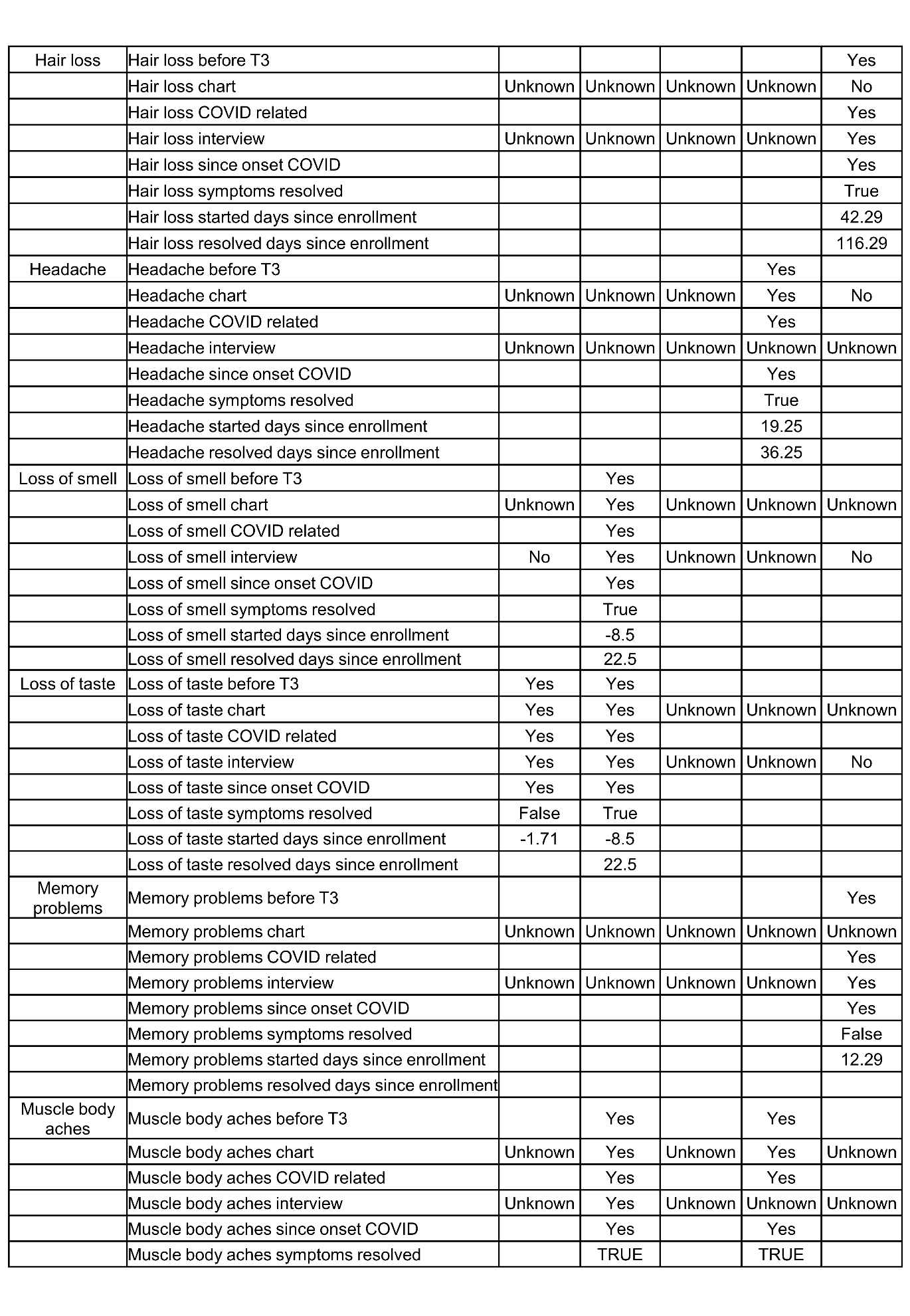

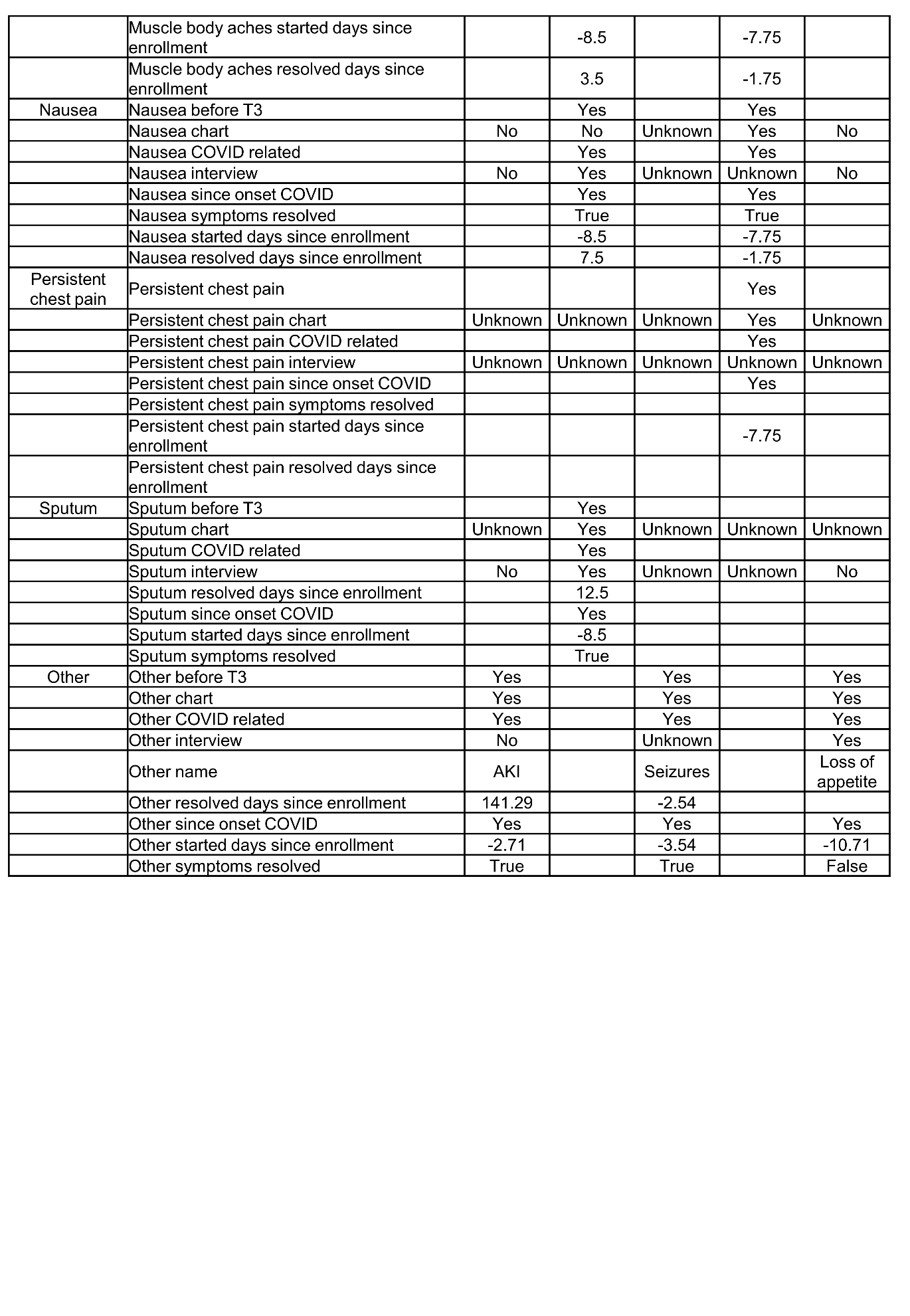

**Table S10: List of MB designs.**

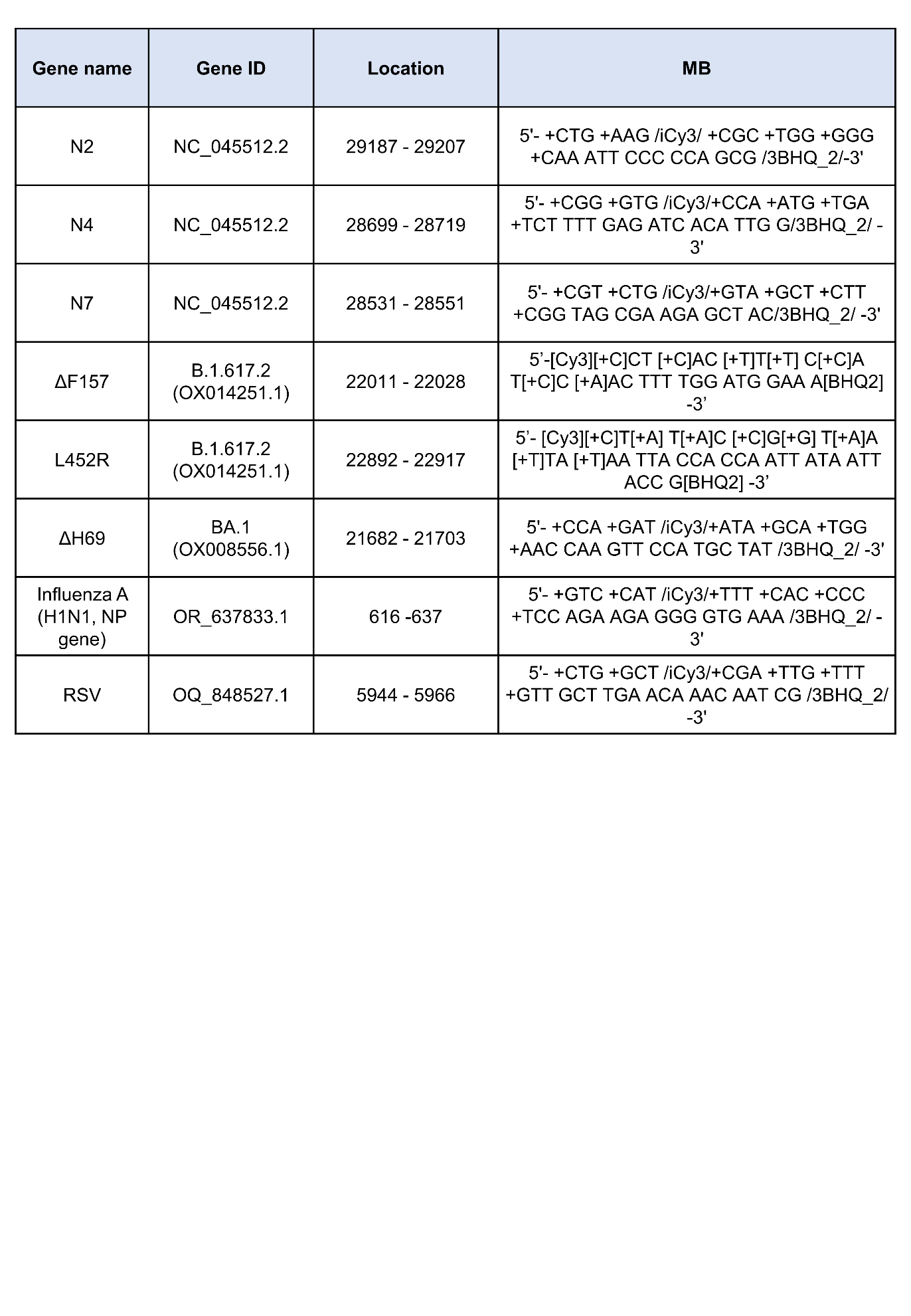
